# Mapping the epigenomic landscape of post-traumatic stress disorder in human cortical neurons

**DOI:** 10.1101/2024.10.11.24315258

**Authors:** Diana L. Núñez-Ríos, Sheila T. Nagamatsu, Jose Jaime Martínez-Magaña, Yasmin Hurd, Gregory Rompala, John H. Krystal, Traumatic Stress Brain Research Group, Janitza L. Montalvo-Ortiz

## Abstract

Most epigenetic research on post-traumatic stress disorder (PTSD) has primarily focused on DNA methylation (5mC) in peripheral tissues, particularly at CpG sites. DNA hydroxymethylation (5hmC) has been found to be highly enriched in the mammalian brain, while 5mC at non-CpG sites shows high enrichment in neurons. However, little is known about their role in PTSD. Here, we characterize genome-wide differential 5mC and 5hmC at both CpG and non-CpG sites in postmortem orbitofrontal neurons from PTSD cases and controls. Utilizing reduced-representation oxidative bisulfite sequencing, we found that genome-wide significant (GWS) differential CpGs were primarily hyper-5mC/5hmC, whereas GWS differential non-CpGs were hypo-5mC/5hmC. Compared with 5mC, we show that 5hmC is a more sensitive epigenetic mark in PTSD, with a higher number of differential 5hmC sites and a stronger significance in enriched pathways. Integrating other -omics data highlighted developmental processes as significant convergent pathways and revealed overlap of our GWS 5hmC findings with 50 previously reported PTSD-associated genes, including potential therapeutic targets such as CRHR1 and DRD4. This study underscores the importance of evaluating 5hmC in the human brain and our multi-omics integration provides insights into potential target genes for future therapeutic interventions in PTSD.

**Graphical abstract:** The study conducted a comprehensive genome-wide analysis of differential 5mC and 5hmC modifications at both CpG and non-CpG sites in postmortem orbitofrontal neurons from 25 PTSD cases and 13 healthy controls. It was observed that PTSD patients exhibit a greater number of differential 5hmC sites compared to 5mC sites. Specifically, individuals with PTSD tend to show hyper-5mC/5hmC at CpG sites, particularly within CpG islands and promoter regions, and hypo-5mC/5hmC at non-CpG sites, especially within intragenic regions. Functional enrichment analysis indicated distinct yet interconnected roles for 5mC and 5hmC in PTSD. The 5mC marks primarily regulate cell-cell adhesion processes, whereas 5hmC marks are involved in embryonic morphogenesis and cell fate commitment. By integrating published PTSD findings from central and peripheral tissues through multi-omics approaches, several biological mechanisms were prioritized, including developmental processes, HPA axis regulation, and immune responses. Based on the consistent enrichment in developmental processes, we hypothesize that if epigenetic changes occur during early developmental stages, they may increase the risk of developing PTSD following trauma exposure. Conversely, if these epigenetic changes occur in adulthood, they may influence neuronal apoptosis and survival mechanisms.

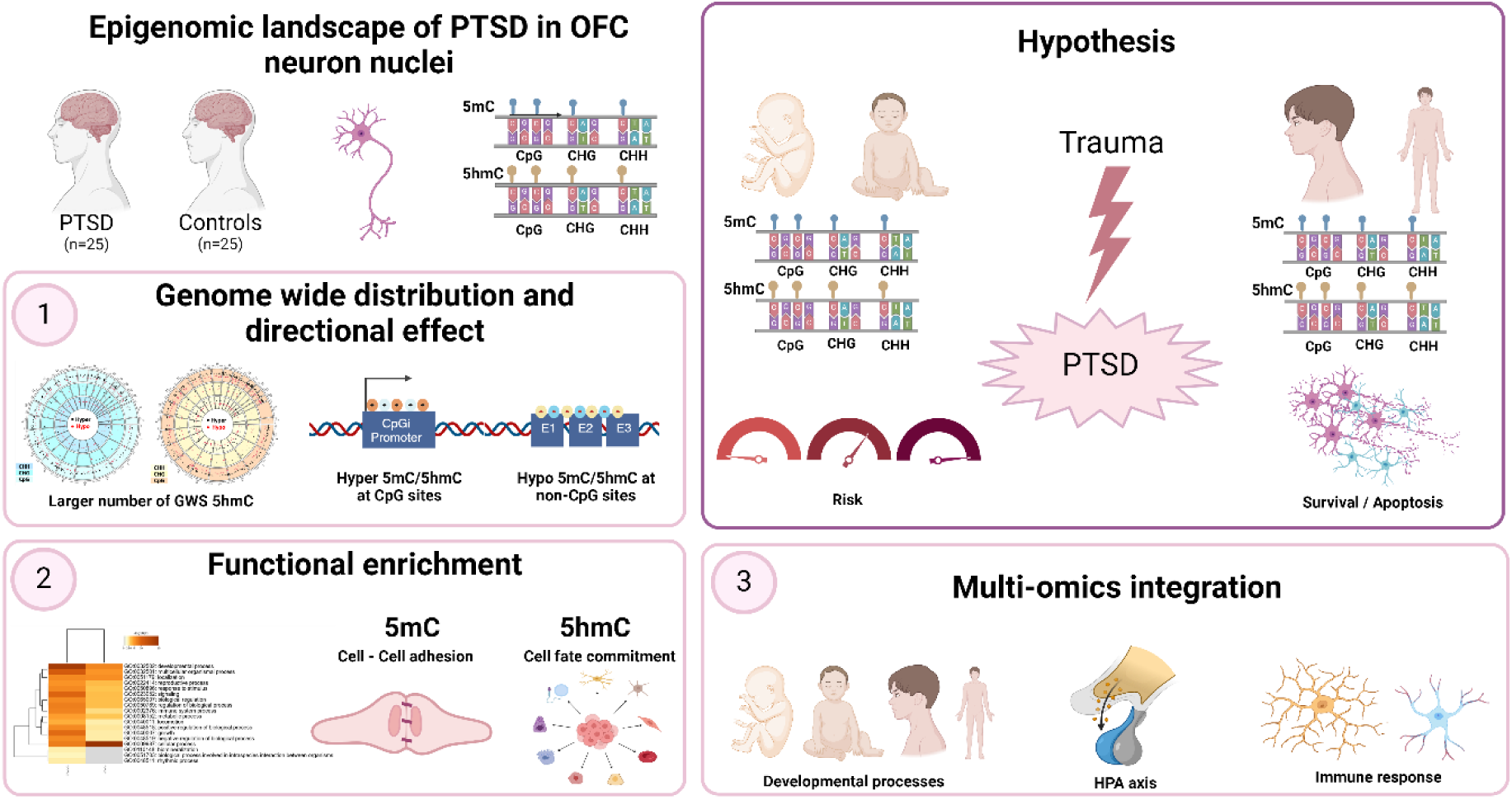

## Introduction

Posttraumatic stress disorder (PTSD) is a complex psychiatric disorder that can develop in individuals exposed to traumatic events (DSM-5) [1]. While nearly 50% of individuals in the U.S. have experienced at least one traumatic event during their lifetime [2–5], the prevalence of PTSD in the general population is approximately 8.3% [6]. The role of genetic variants in PTSD has been revealed in recent large-scale genome-wide association studies (GWAS) reporting a SNP-based heritability of 5-20% [7,8]. Multiple environmental factors are also known to play a role in PTSD risk [7,9].

Epigenetic mechanisms have been suggested to underlie the gene-environment interplay in PTSD risk. One of the most commonly studied epigenetic mechanisms in humans is DNA methylation (5mC) at CpG sites (CpGs), characterized by the addition of a methyl group to the 5’ position of a cytosine ring linked to a guanine [10]. Epigenome-wide association analyses (EWAS) from our group and others have revealed multiple epigenetically-dysregulated genes in PTSD [11–16]. Genes such as *AKT, ANK3, BDNF, CNR1, COMT, CREB, DRD2, DMRTA2, DOCK2, EFS, ELK1, ETS-2,* and *GATA3* have been associated with PTSD in EWAS studies evaluating peripheral samples (blood and saliva) [11–16]. Notably, studies evaluating 5mC in the brain at the genome-wide level have been primarily limited to animal models, in which differentially methylated genes have been involved in inflammation, neurogenesis, and synaptic plasticity [17–19]. Considering the tissue- and cell-type specificity of epigenetic patterns [20] and the known impact of PTSD on the brain, human studies evaluating brain tissue are greatly warranted.

Beyond the existing gap in 5mC patterns in the brain, there are other DNA-epigenetic mechanisms highly enriched in the brain with distinct gene regulatory processes that should be studied. One such mechanism is DNA hydroxymethylation (5hmC), an oxidized 5mC that represents a stable epigenetic mark and part of the demethylation pathway [21–25]. 5hmC is often co-localized with enhancer elements [26,27] and modulates the binding of DNA glycosylases and DNA repair proteins to DNA [28,29]. Promoter and gene body regions of actively transcribed genes are often enriched by 5hmC marks [25]. Studies have linked 5hmC to psychiatric disorders including depression [30], alcohol use disorder [31,32], and, more recently, opioid use disorder [33], where functional differences between 5hmC and 5mC are postulated. To our knowledge, no studies have evaluated the role of 5hmC in PTSD.

Additional DNA epigenetic marks within the human brain that warrant further investigation is 5mC at non-CpG sites (CHH and CHG, where H refers to 5mC at cytosines linked to adenine, thymine, or other cytosines) [34,35]. 5mC at non-CpGs is reported in a limited number of cell types such as stem cells, oocytes, neurons, and glia. Indeed, neurons exhibit a distinct non-CpG 5mC pattern compared to glial cells, highlighting cell-type specificity. Non-CpG 5mC accumulates throughout development, particularly increasing during synaptogenesis and synaptic pruning. Additionally, 5mC at non-CpGs is known to play a critical role in cognitive function [34–38]. Very few epigenetic studies have evaluated 5mC at non-CpGs in the context of psychiatric disorders. Evidence of the role of 5mC at non-CpGs in opioid use disorder (OUD) was recently reported by our group [39]. To our knowledge, no studies have evaluated the role of 5mC and 5hmC at non-CpG sites in PTSD.

The orbitofrontal cortex (OFC) is a brain region implicated in emotion regulation and fear response, both of which are critical domains in PTSD [40–43]. Here, we present the first neuronal-specific mapping of 5mC and 5hmC at CpGs and non-CpGs in the human postmortem OFC of individuals with PTSD and controls (**Fig 1**). We reveal differences and similarities between 5mC/5hmC at CpG and non-CpG sites in terms of genome distribution and directional effect and demonstrate that 5hmC emerges as a prominent epigenetic marker in PTSD. We integrate our epigenomic findings with previously reported findings from other-omic studies in PTSD (i.e., GWAS, EWAS, and transcriptomics) to explore convergence on molecular mechanisms and prioritize dysregulated genes and gene networks that may play an essential role in PTSD. We found that developmental processes were a highly enriched mechanism in PTSD, and more than 50 PTSD-associated genes were prioritized through multi-omics analysis. Lastly, we conduct a drug-repurposing analysis and report potential therapeutic targets for the treatment of PTSD such as *CRHR1* and *DRD4*.

**Fig. 1.**
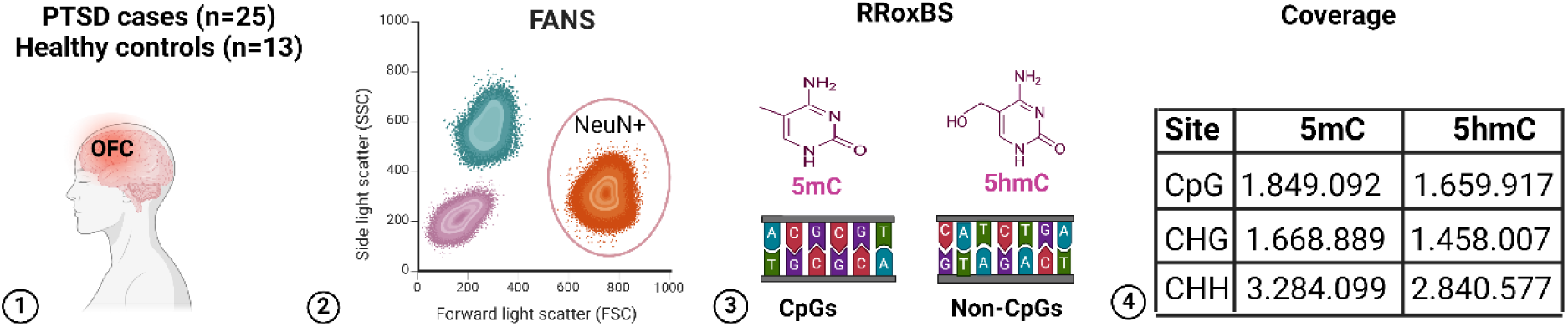
Study workflow. 1. Postmortem brain tissue from the orbitofrontal cortex (OFC) was obtained from individuals with PTSD and healthy controls collected at the VA’s National PTSD Brain Bank. 2. NeuN+ nuclei were isolated through FANS (fluorescence-activated nuclei sorting). 3. Reduced-representation oxidative-bisulfite-sequencing (RRoxBS) was conducted to assess 5mC and 5hmC in CpG and non-CpG sites (i.e., CHG/CHH). 4. The coverage of the RRoxBS is described, including the number of CpG, CHG, and CHH sites for 5mC and 5hmC.

## Results

### 1. Differential DNA methylation at CpG and non-CpG sites

We identified 735 genome-wide significant (GWS; q-value < 0.05 and ± Δ value higher than 2%) differential 5mC sites associated with PTSD, including 539 at CpGs and 196 at non-CpGs (**Fig. 2a and Table S1**). Notably, the directional effect of GWS 5mC sites varied between CpGs and non-CpGs. While 64.56% of CpG sites were hypermethylated (higher methylation in PTSD individuals as compared to healthy controls), on average, 64.03% of non-CpGs (50.59% at CHG sequence and 77.48% at CHH sequence) exhibited hypomethylation (**Table S1**). Regarding genome-location mapping of the GWS 5mC sites, 43.23% of CpGs and 33.90% of non-CpGs (24.72% at CHG and 18.02 at CHH) map to promoter regions (**Fig. 2b**). GWS 5mC sites mapping CpG islands correspond to 43.41% of CpGs and 22.85% of non-CpGs (25.88% at CHG and 19.82% at CHH) (**Fig. 2c**). For the GWS 5mC sites mapping CpG islands within promoter regions, we observed hypermethylation in 76.67% of CpGs and hypomethylation in 93.33% of non-CpGs. These results show that both GWS differential 5mC CpGs and non-CpGs associated with PTSD map to important regulatory regions involved in gene expression.

**Fig. 2.**
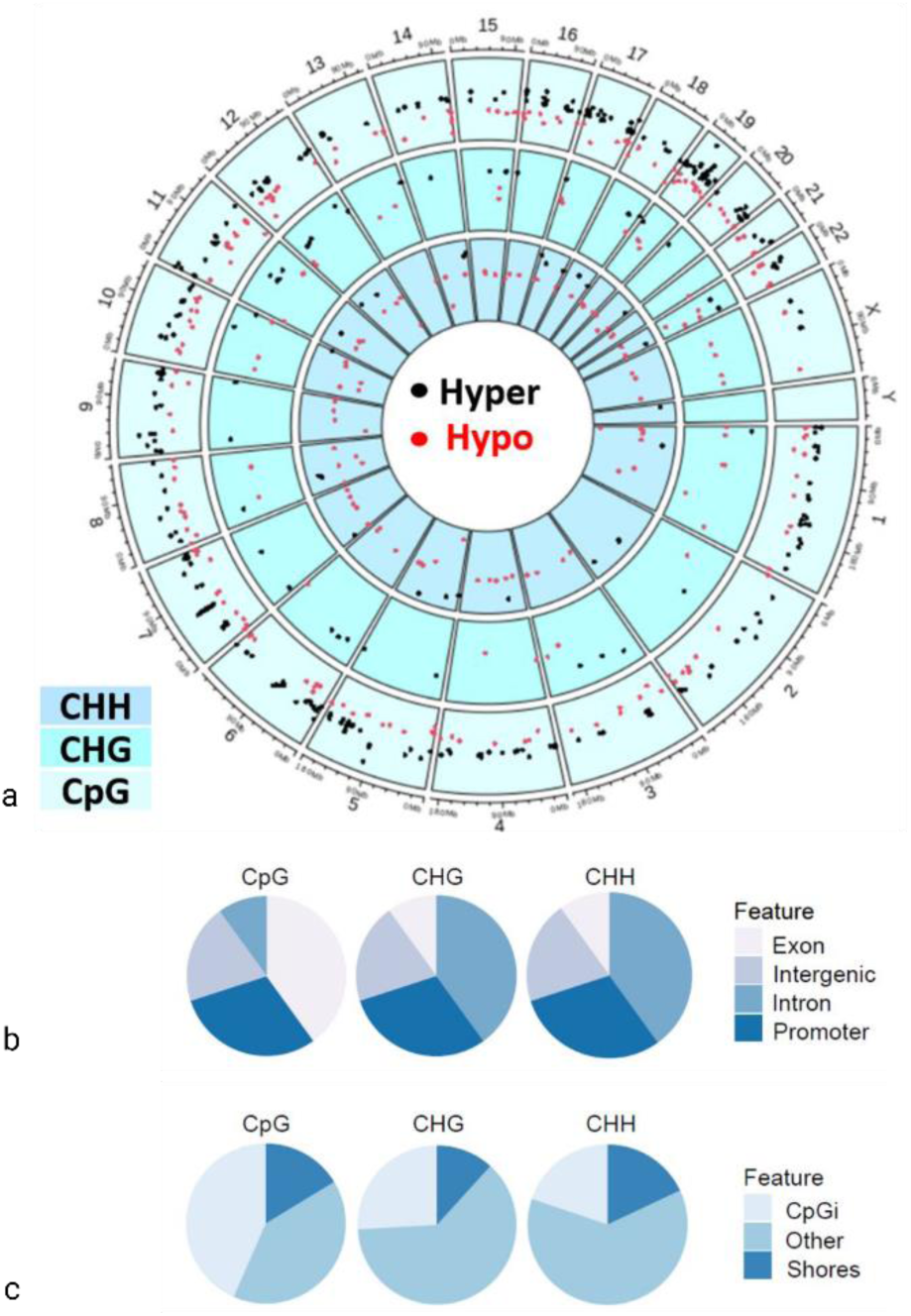
PTSD-associated significant differentially-methylated cytosines in OFC neuronal nuclei. A) Circos plot shows the genome-wide distribution of differentially-methylated cytosines at GWS CpGs and non-CpG sites associated with PTSD. b) and c) Pie charts representing gene and CpGi features of significant genome-wide significant CpG and non-CpGs.

To explore the molecular mechanisms that may be impacted by 5mC alterations associated with PTSD, we performed a functional-enrichment analysis. After multiple-testing correction (Q value < 0.05), biological pathways such as developmental process (GO:0032502) and response to stimulus (GO:0050896) were significantly enriched by both GWS 5mC CpGs and non-CpGs (**Fig. S1 and Tables S2-3**). In the co-methylation analysis for CpGs, we identified 16 modules with a correlation score greater than 0.4 with PTSD (six with positive correlation and ten with negative correlation) (**Fig. S2a-c**). Brain development (GO:0007420) was a common pathway enriched in the thistle3, olivedrab2, and yellowgreen modules (**Fig. S2d and Table S4**). For non-CpGs (correlation score higher than 0.2), one module for CHG sites was negatively correlated with PTSD, showing enrichment for cellular response to retinoic acid (GO:0071300) (**Fig. S2a-d**).

Overall, our PTSD-associated 5mC findings in the OFC neuronal nuclei reveal distinct patterns of regulatory profiles when comparing CpGs with non-CpGs, though both affect similar biological pathways such as developmental processes and response to stimulus.

### 2. Differential DNA hydroxymethylation at CpG and non-CpG sites

Compared to GWS 5mC findings, a higher number of GWS-differential 5hmC sites was associated with PTSD, including 816 at CpGs and 334 at non-CpGs (103 CHGs and 231 CHHs) (**Fig. 3a and Table S5**). As with 5mC, a higher number of GWS 5hmC sites were observed in CpGs when compared to non-CpGs. Hyper-5hmC was identified in 58.82% of CpGs and hypo-5hmC in 64.62% of non-CpGs (62.14% at CHG and 67.10% at CHH) (**Table S5**). Gene-feature annotation was also similar to 5mC, with 46.57% of GWS 5hmC at CpGs and 27.12% at non-CpGs mapping to promoters (29.13% in CHG and 25.11% in CHH sites) (**Fig. 3b**). The percentage of GWS-differential sites mapping CpG islands was slightly higher in 5hmC CpGs compared to 5mC: 56.50% in CpGs and 22.85% in non-CpGs (25.88% in CHG and 19.82 in CHH sites) (**Fig. 3c**). Regarding GWS 5hmC sites mapping CpG islands within promoter regions, hyper-5hmC was observed in 68.48% of CpGs and hypo-5hmC in 88.57% of non-CpGs. Overall, the directions of effect and genomic locations of the PTSD-associated GWS 5hmC resemble the pattern observed in 5mC, both mapping gene regulatory regions.

**Fig. 3.**
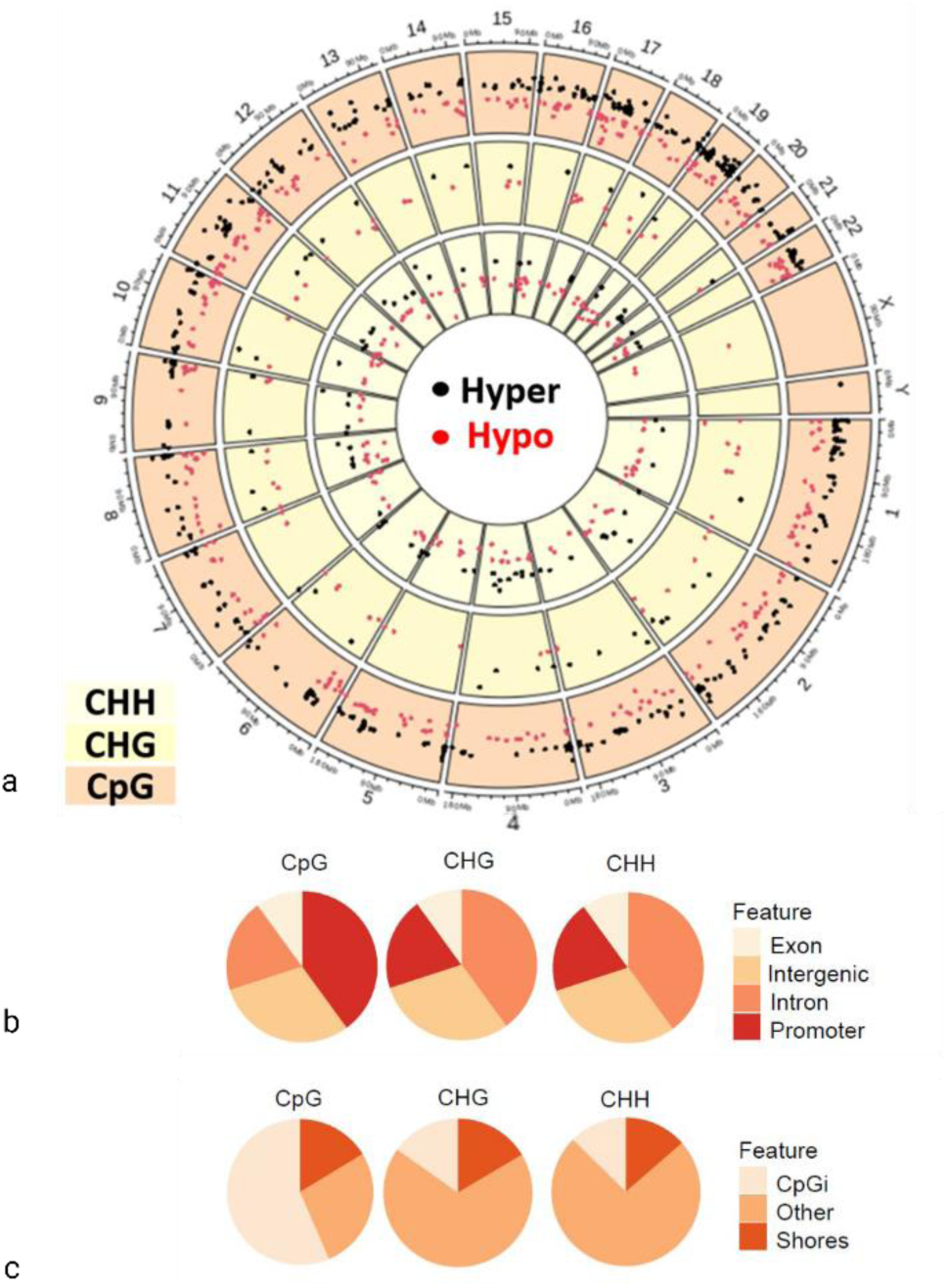
PTSD-associated significant differentially-hydroxymethylated cytosines in OFC neuronal nuclei. a) Circos plot shows the genome-wide distribution of significantly differentially-hydroxymethylated cytosines at CpG and non-CpG sites associated with PTSD. b) and c) Pie charts depict the gene and CpGi features of significant CpGs and non-CpGs.

At the functional level, the enrichment for developmental process (GO:0032502) and response to stimulus (GO:0050896) observed in GWS-differential 5mC sites was also found in GWS-differential 5hmC sites, although at a greater significance. Additional biological pathways such as signaling (GO:0023052), cellular process (GO:0009987), and metabolic process (GO:0008152) were also significantly enriched in GWS- differential 5hmC sites (**Fig. S3 and Tables S6-7**). For the co-hydroxymethylation at CpGs, ten modules were associated with PTSD (**Fig. S4a-d**) and significantly enriched for cell morphogenesis (GO:0000902), head development (GO:0060322), immunoglobulin production (GO:0002377), and lymphocyte apoptotic process (GO:0070227) (**Fig. S4d and Table S8**). Co-hydroxymethylation analysis at non-CpGs identified the mediumaquamarine module as significantly associated with PTSD for CHH sites (**Fig. S4a-d**), showing enrichment for homophilic cell adhesion via plasma membrane adhesion molecules (GO:0007156) and embryo development ending in birth or egg hatching (GO:0009792) (**Fig. S4d and Table S8**). For CHG sites, we identified the mistyrose2 module, but no significant functional enrichment was found.

While our findings reveal a notable similarity between the 5mC and 5hmC patterns associated with PTSD, the higher frequency of GWS 5hmC sites linked to PTSD, along with their higher enrichment in critical biological processes, underscores the pivotal role of 5hmC in the context of PTSD.

### 3. Developmental processes as a convergent enrichment pathway

Next, we explored convergent pathways across all domains, including 5mC and 5hmC, as well as CpG and non-CpG sites (**Tables S9-10**), to better understand the epigenetic landscape of PTSD in human cortical neurons. When comparing genes mapping GWS-differential 5mC and GWS 5hmC sites (in either CpGs or non-CpGs), we observed convergent enrichment in cellular processes (GO:0009987) and developmental process (GO:0032502). Within cellular processes, enrichment in homophilic cell adhesion via plasma membrane adhesion molecules (GO:0007156) and cell-cell adhesion via plasma membrane adhesion molecules (GO:0098742) exhibited a higher significance in GWS-differential 5mC findings. The enrichment in developmental processes was primarily driven by GWS-differential 5hmC, including embryonic morphogenesis (GO:0048598), cell-fate commitment (GO:0045165), and neuron-projection development (GO:0031175) (**Fig. 4a-b**). Comparing GWS-differential CpG and non-CpG sites for either 5mC or 5hmC, we observed that the enrichment of embryonic morphogenesis (GO:0048598), cell-fate commitment (GO:0045165), and neuron-fate commitment (GO:0048663) (**Fig. S5**) was predominantly influenced by differential 5hmC CpGs. We showed that enrichment in developmental processes was primarily driven by GWS-differential 5hmC in CpG sites.

**Fig. 4.**
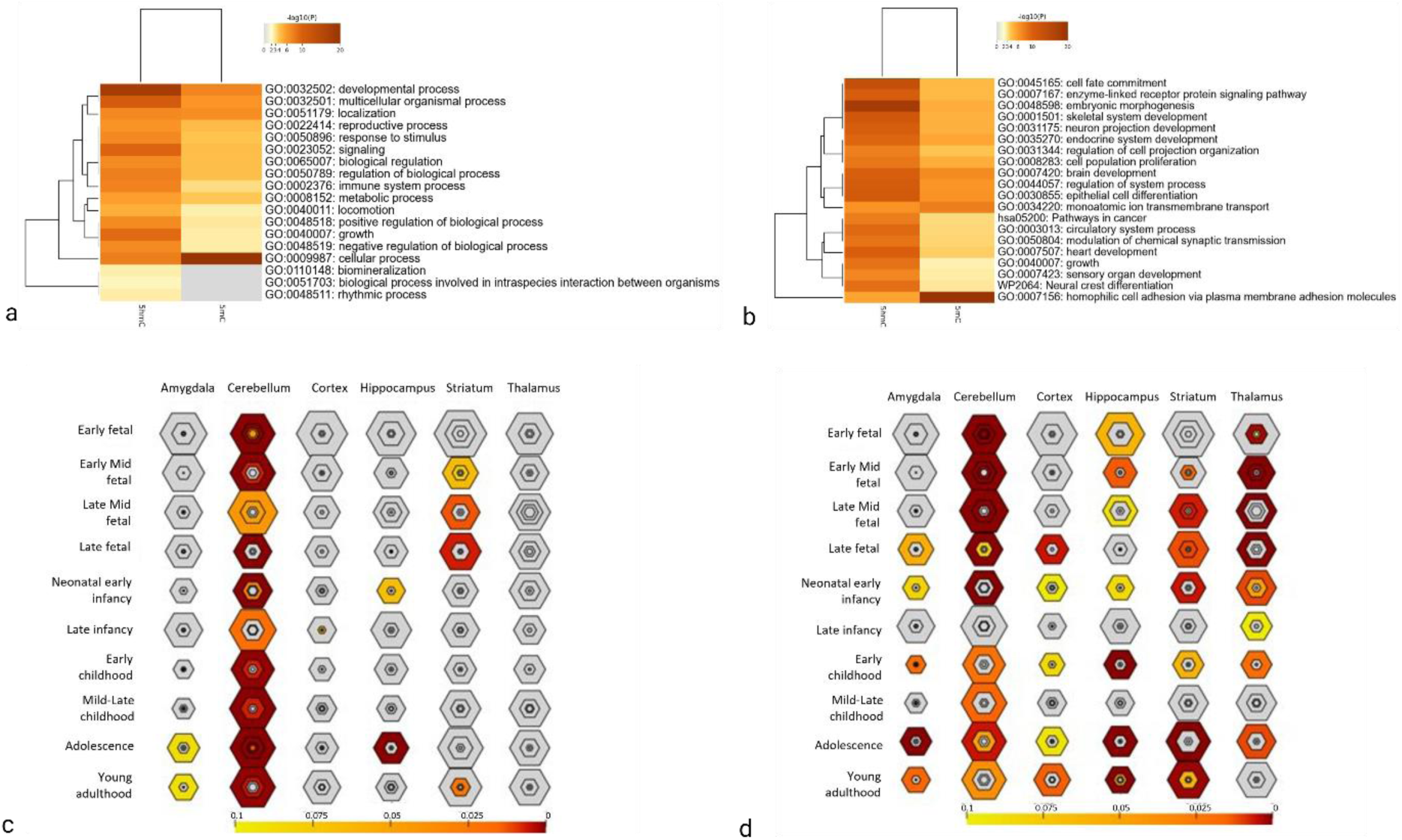
Convergent pathways for neuronal-specific PTSD-associated epigenomic changes in the OFC. a) Enriched GO parental pathways by epigenetic mark (5mC/5hmC). b) Enriched GO pathways by epigenetic mark (5mC/5hmC). c) Gene-set enrichment analyses for genome-wide significant differential 5mC across brain-developmental stages. d) Gene-set enrichment analyses for genome-wide significant differential 5hmC across brain-developmental stages.

Given the observed enrichment of PTSD-associated epigenetic marks in developmental processes, we explored their enrichment across various developmental stages and different brain regions (**Tables S11- 12**). In the brain cortex, GWS-differential 5hmC marks showed a significant enrichment for late fetal, early infancy, early childhood, and adulthood stages (p value < 0.05; **Fig. 4c-d and Table 1**). In terms of brain regions, GWS-5hmC marks were enriched in the striatum, thalamus, hippocampus, and amygdala (**Fig. 4c-d and Table S12**). The highest brain-region enrichment for both GWS-differential 5mC and 5hmC sites across several developmental stages was the cerebellum (**Fig. 4c-d and Table S11-12**).

**Table 1.** GWS-differential 5mC and 5hmC sites associated with PTSD enriched in the brain cortex across different developmental stages.

|  | Methylation | Hydroxymethylation |
| --- | --- | --- |
| Late fetal | *SDK1, RTN4RL1, PCDHAC1, EDARADD, PCDHA5, PCDHA7, PCDHA3, PCDHA4, PCDHA2, SLCO2A1, ULBP2, PCDHA13, NFE2L3, CSMD2, COL15A1, UST, STK31, OCA2, MN1, ADAMTSL3, CPVL, CD1D, SCUBE1, CUX2, COMTD1, and FBLIM1* | *PRSS12, CRIP2, RTN4RL1, THSD7B, PCDHA5, TBXA2R, PCDHA4, TBC1D30, PCDHA2, TNFAIP2, PTPN2, PCDHA3, MEPE, TACR3, VSTM2L, SMTNL2, COMTD1, TRPC7, CSMD2, COL15A1, ARHGAP15, UST, OCA2, MN1, ADAMTSL3, CHST8, SCUBE1, CUX2, EPS8L1, C1QL1, KLHL8, TMEM102, and PTPRK* |
| Early infancy | *ULBP2, F7, UST, PCDHA5, STK31, TWIST2, RTN4RL1, SHANK2, CHRM1, PCDHA9, NEURL1B, ST14, DLGAP2, CD1D, CPLX3, LRRK1, TMEM132B, LINGO1, CUX2, and KALRN* | *F7, FAM43A, TWIST2, RTN4RL1, PCDHA5, KALRN, CPLX3, LRRK1, RXFP3, MEPE, LINGO1, VSTM2L, SDK2, CHRM1, GP1BB, ST14, LAMC3, GRIN1, UST, FAM163B, DLGAP2, MATK, CUX2, SHANK2, CHST13, and TMEM151B* |
| Early childhood |  | *MERTK, SCT, CDC42EP5, PPP1R3C, HHIPL1, EPAS1, TWIST2, ISG20, SOX17, FAM163B, SMAD6, DLGAP2, SCARA3, MATK, GAS2L1, KALRN, ZNF366, and GUK1* |
| Adulthood |  | *F7, MAPK8IP3, TWIST2, JAKMIP3, KALRN, PTH2R, LRRK1, CORO6, TNFRSF11A, FBXL16, SCN8A, PLEKHA2, CHRM1, UNC13A, GRIN1, VWA5B2, PPP2R2C, BTBD9, TRPM2, FAM163B, ATP6V1B2, DLGAP2, KCNA1, GDF7, SLC12A5, C1QL1, FGF8, FAM153B, MEPE, PTPRK, MCF2L2, PDE4A, and UNC5A* |

We also conducted a disease-enrichment analysis on genes mapping to the GWS-differential sites associated with PTSD (**Tables S13-16**). The highest enrichments observed for GWS-differential 5mC and 5hmC sites were vital capacity (C0042834) and diastolic blood pressure (C0428883), respectively (**Tables S13-16**). While mental disorder was enriched for both GWS-differential 5mC and 5hmC genes, a higher enrichment was observed with 5hmC, particularly at CpGs (**Fig. 5**). All GWS-differential 5mC and 5hmC sites mapping to genes enriched in mental disorders exhibit predicted transcription-factor binding sites (TFBS) (**Table 2**), suggesting a potential crucial role in gene-transcription regulation. Interestingly, a significant enrichment for PTSD was also observed, with *CHRNA4, CRHR1, DRD4, ESR1, FLT4, HSP90AA1, INSRR, LMNA, LRP1, LTBP3, NOTCH3, OPRL1, PDE4A, PPP1R3C, TP73, TPH1, KL, HDAC4, C1QL1, PDAP1,* and *IMPACT* mapping to GWS differential 5hmC sites, while *NPY and TERT* mapped to both GWS-differential 5mC and 5hmC sites (**Table S13-16**).

**Fig. 5.**
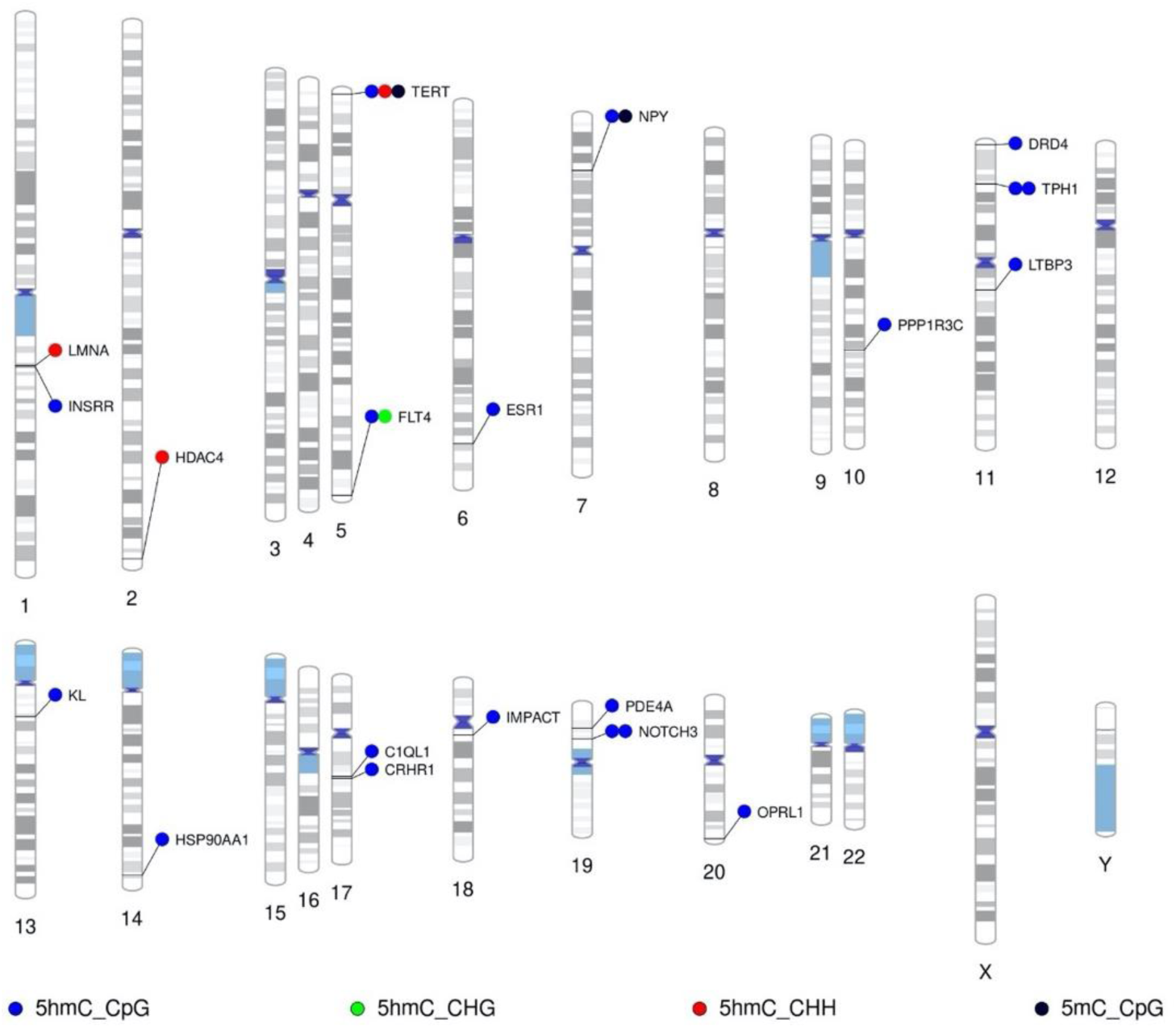
PTSD-associated epigenomic alterations in OFC neuronal nuclei are enriched for mental disorders. Ideogram depicting PTSD-associated genome-wide hydroxymethylated genes at CpG and non-CpG sites enriched for mental disorders.

**Table 2.**
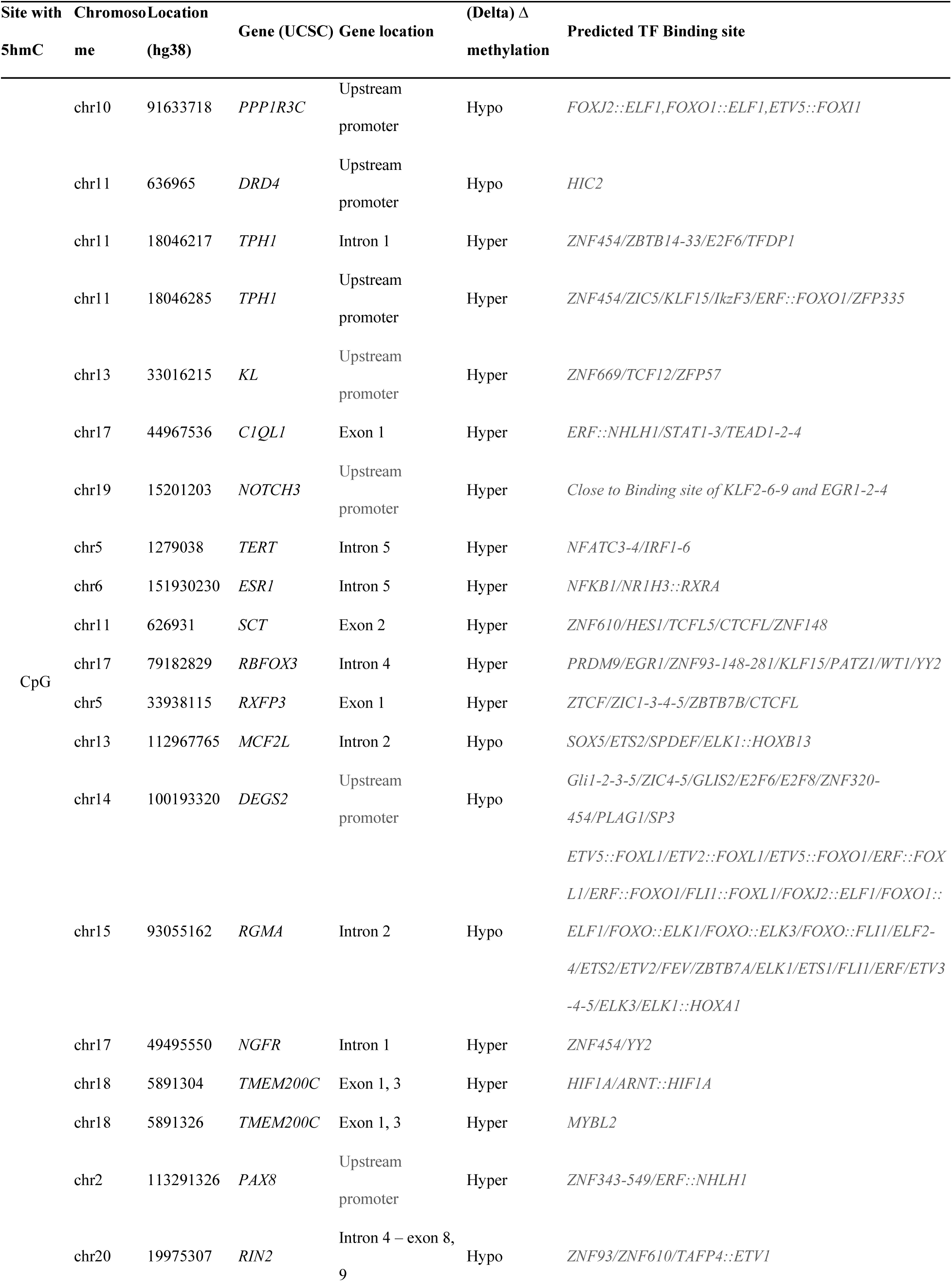
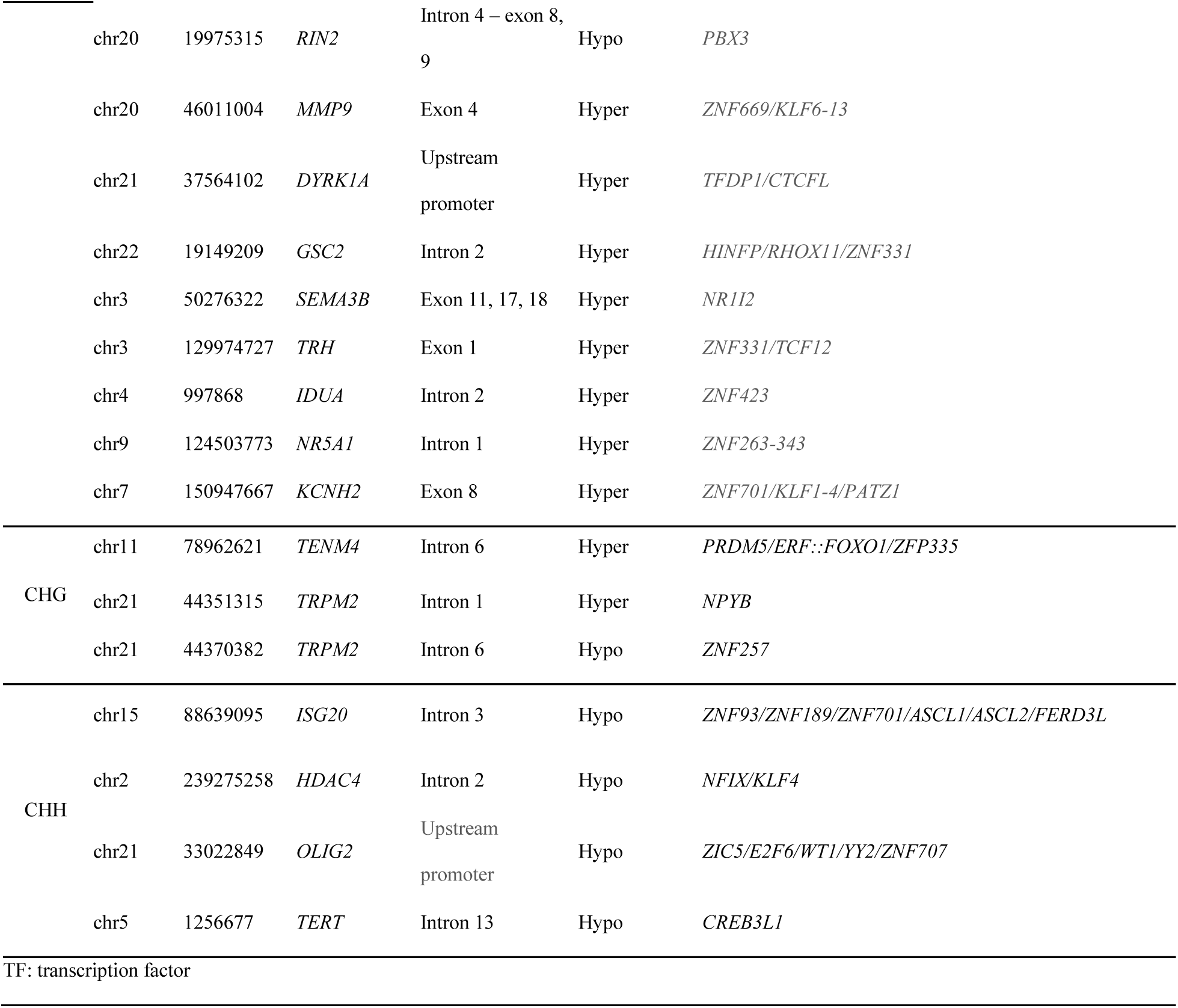
Enriched genes for mental disorders with predicted transcription-factor (TF) binding sites.

Collectively, our GWS-differential 5mC and 5hmC findings showed a prominent role for hydroxymethylation in PTSD, particularly in CpGs of genes involved in developmental processes. Furthermore, genes enriched in mental disorders and PTSD primarily exhibited GWS-differential 5hmC at CpGs.

### 4. Convergent multi-omics findings

We compared our current findings with prior-omics studies of PTSD, examining their convergence at gene and pathway levels in central and peripheral tissues, to help prioritize PTSD-relevant genes and pathways.

#### a. Convergence in OFC

To prioritize genes and molecular pathways altered in the brains of individuals with PTSD, we analyzed the convergence of our findings with previously reported-omics studies obtained from central tissues. Specifically, we examined the convergence between our neuronal-specific OFC epigenomic findings with transcriptomic findings obtained from bulk-OFC [9].

At the gene level, our GWS-differential 5mC and 5hmC findings, along with the published PTSD transcriptomic findings [9], reveal an overlap of five genes: *RTN4RL1, RASAL3, BIN2, FOXO3,* and *ITGAX* (**Table S17**). *RTN4RL1*, upregulated in the bulk-OFC, exhibited one hyper-5mC (chr17:1936219) and one hypo-5hmC (chr17:1998713) CpG site in intron 1, along with one hyper-5mC CHH site in exon 2. The GWS-differential 5hmC CpG site in intron 1 of *RTN4RL1* has a predicted transcription-factor binding site (TFBS) for *ZNF148, ZNF281, ZNF454, ZNF610, KLF15, KLF17, PATZ1, MAZ,* and *SP1*. Additionally, the GWS-differential 5mC CHH site in exon 2 had a predicted TFBS for *PRDM9*. *BIN2*, found to be downregulated in the bulk-OFC, was identified with two GWS-differential 5hmC CpGs. One CpG site was hypo-5hmC (chr12:51324364) and located in intron 1 with an H3K27Ac mark and predicted TFBS for *ZBED2*, *IRF6,* and *ZFX*. The second 5hmC CpG site in *BIN2* was hyper-5hmC (chr12:51324346) in intron 1 with an H3K27Ac mark and no predicted TFBS. *RASAL3*, downregulated in the bulk-OFC and dlPFC, was identified with a GWS 5hmC CpG site and one GWS 5mC CHH site. The observed GWS CpG at the *RASAL3* gene corresponds to a hyper-5hmC site (chr19:15457531) located in exon 9 with predicted TFBS for *TCF3*, *TCF4, TCF12*, and *ASCL1*. The observed GWS CHH site at the *RASAL3* gene corresponds to a hypo-5mC site (chr19: 15471745) located upstream of the promoter region, with predicted TFBS for *KLF3* and *CREB3L1*. *FOXO3* was found to be upregulated in the bulk-OFC and identified with a hyper-5hmC CpG site (chr6:108562555) located in intron 1, with an H3K27Ac mark and predicted TFBS for *ZNF93, ZNF708, ZNF189, ZNF610, PRDM9,* and *TFAP4:ETV1*. Lastly, *ITGAX* was found to be downregulated in the bulk-OFC (and dlPFC) and exhibited hyper-5hmC at a CHH site (chr16:31361425) located in intron 9, with a predicted TFBS for *ZNF263 and SP1*.

Regarding convergent molecular pathways and biological processes, our GWS-differential 5mC and 5hmC sites in OFC neuronal nuclei converge with GWS-differentially-expressed genes in the bulk-OFC in developmental processes (GO:0032502) including tube morphogenesis (GO:0035239), blood vessel morphogenesis (GO:0048514), and neuron-projection development (GO:0031175) (**Fig. S6 and Table S18**).

A protein-protein interaction (PPI) network analysis identified focal adhesion, with PI3K-Akt-mTOR-signaling pathway (WP3932) and assembly and cell-surface presentation of NMDA receptors (R- HSA-9609736) among the convergent mechanisms across the epigenetic and transcriptomic results in OFC (**Fig. 6**). Overall, convergent analysis in the human OFC highlights developmental processes as a key biological pathway in PTSD.

**Fig. 6.**
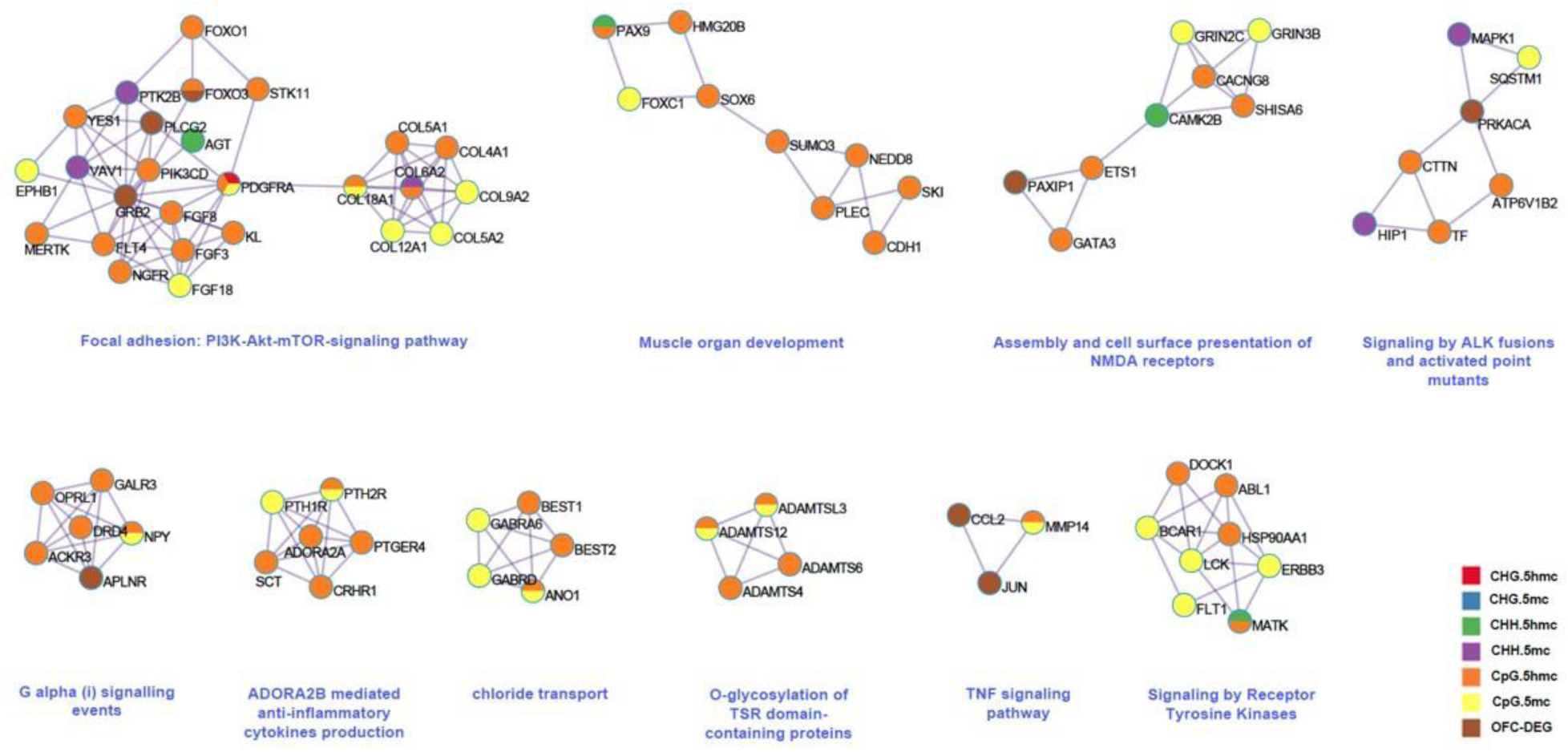
Cross-tissue multi-omics convergence in PTSD. Protein-protein interaction networks where PTSD-associated OFC findings converge, integrating 5mC and 5hmC and CpG, and CHH, together with differentially expressed genes in the OFC.

#### b. Cross-tissue, Multi-omics Convergence

For the multi-omics integrative analysis, we evaluated convergence across genomic, epigenomic, and transcriptomic data by integrating results from both central and peripheral tissues. At the gene level, a significant overlap was found by performing a Fisher test (OR=2.5, *p*-value 3.6×10^-8^). Our PTSD-associated GWS-differential 5mC and 5hmC findings overlapped with 50 PTSD-associated genes previously reported in GWAS, EWAS, or transcriptomic studies conducted in human peripheral and brain tissues. There were ten genes with differential 5mC (*PHTF1, PCDHA1, GLIS3, SOX6, SIPA1, NACA, MNT, SDK1, NINJ2,* and *POGK*) and 40 genes with differential 5hmC (*CRHR1, NRXN1, PCDHA1, TSNARE1, PGPEP1, PIK3CD, RNF220, NENF, BEST1, CRIP1, MAF, MED9, LIMD2, ZADH2, CDC37, NCL, OPRL1, TRIOBP, PLXNB2, CHST13, RGS12, TTYH3, EPHB4, ANK1, ATP2A3, SMAD7, COPA, BRI3BP, ARHGAP15, HDAC4, GSK3B, PIP5K1C, DHX37, VAMP2, AHRR, DUSP22, TRPS1, NDRG1, UST,* and *GRIN1*) (**Table S17**).

Localization (GO:0051179), cellular process (GO:0009987), and regulation of biological process (GO:0050789) were among the parental pathways enriched across all –omics studies (**Fig S7**). Particularly, cell-cell adhesion (GO:0098609), PID PDGFRB pathway (M186), B-cell-receptor signaling pathway (WP23), regulation of monoatomic ion transmembrane transport (GO:0034765), regulation of trans-synaptic signaling (GO:0099177) and neuron-projection development (GO:0031175) are convergent pathways across previously reported epigenomic, transcriptomic, and GWAS PTSD-associated findings (**Table S19**). Cross-tissue epigenetic findings associated with PTSD converged in pathways such as receptor tyrosine kinases (R-HSA-9006934), gland development (GO:0048732), regulation of secretion (GO:0051046) and regulation of the MAPK cascade (GO:0043408). Overall, our cross-tissue multi-omics convergent analysis underscores 5hmC as the primary epigenetic mark that maps genes previously associated with PTSD.

### 5. Drug discovery

A drug-repurposing analysis was conducted to evaluate the enrichment of known drugs interacting with genes containing GWS-differential 5mC/5hmC sites associated with PTSD. We found that 37 genes have a significant interaction with certain known drugs. These included *PDGFRA* for GWS-differential 5mC sites, *VAMP2, LTB4R, ALOX5, MAP3K13, KCNN3, INSRR, DRD4, BRSK2, TPH1, SLC6A5, P2RY6, MLNR, DLL4, RGMA, RARA, FZD2, CRHR1, MADCAM1, TBXA2R, PDE4A, NOTCH3, ACKR3, MMP9, SLC12A5, OPRL1, TRPM2, ADORA2A, TACR3, LVRN, TRPC7, KCNK17, ABL1, ENTPD2,* and *CACNA1B* for GWS-differential 5hmC sites, and *TERT* and *EGLN3* for both GWS-differential 5mC and 5hmC (**Table S20**). Notably, 13 of these 37 genes were found among the genes with GWS-differential 5mC/5hmC sites enriched for mental disorders. Furthermore, several known drugs target reported PTSD-associated genes. For example, verucerfont, ONO-2333MS, pexacerfont, SSR125543, ontamalimab, and antalarmin were identified as antagonists of *CRHR1.* Dexfenfluramine and sibutramine were identified as antagonists of *SLC6A4*. Nadide and econazole were identified as an agonist and an antagonist of *TRPM2*, respectively.

## Discussion

The present study reveals the neuronal methylome and hydroxymethylome profiles associated with PTSD within the human OFC neurons, spanning both CpG and non-CpG sites. Diverse patterns of 5mC/5hmC profiles were found between CpG and non-CpG sites, exhibiting distinct directional effect and genome-wide distribution. We identified a higher number of differential sites with 5hmC than with 5mC marks, along with enrichment in developmental processes, mental disorders, and PTSD driven by 5hmC marks. Furthermore, the highest convergence with previously-reported PTSD-omics findings was observed with genes mapping to 5hmC differential marks. Our findings make a significant contribution by investigating understudied epigenetic mechanisms in PTSD in a cell-type specific manner.

Research indicates that while approximately 60-70% of CpGs are methylated, the CpGs located within promoter-associated CpG islands often exhibit low 5mC levels [35,44,45]. Similarly, 5hmC is depleted in CpG islands and enriched in enhancers, gene bodies, and CpG shores [46,47]. Non-CpG sites exhibit enrichment in gene bodies, particularly at exons [48]. In our study, we observed variation between CpG and non-CpG sites. While GWS-differential CpGs tending towards hyper 5mC/5hmC mainly mapped to promoter regions and CpG islands, GWS-differential marks at non-CpGs showing tendencies towards to hypo-5mC/5hmC mapped to intronic regions and open sea. Our findings suggest that individuals with PTSD exhibit a significant epigenetic dysregulation in OFC neuronal cells, particularly characterized by changes in 5hmC within regulatory regions such as CpG islands and promoters.

Our study also highlights the value of examining both 5mC and 5hmC at non-CpGs. Previous studies have shown that 5mC marks in mammals extend beyond non-CpGs in a cell-type-specific manner. Notably, neuronal cells exhibit higher levels of 5mC at non-CpGs compared to other brain cell types [34,35,49]. In the OFC neuron nuclei, we found a significant amount of GWS-differential 5mC and 5hmC marks associated with PTSD at non-CpG sites. The genome distribution of these non-CpG sites was consistent with that reported in the mammalian brain [35,49]; that is, a reduced co-localization between 5mC CpG and non-CpGs as well as hypo-5mC in non-CpGs proximal to CpGs. However, the GWS- differential 5mC/5hmC non-CpG sites in our study did not exhibit enrichment at exons [48]. Instead, they predominantly mapped to intronic regions, displaying tendencies towards hypo-5mC/5hmC in PTSD cases compared to controls. This divergence underscores the complexity of epigenetic regulation in the context of PTSD, emphasizing the need for further exploration into non-CpG methylation and hydroxymethylation dynamics.

Regarding the potential functional impact of epigenetic findings, it has been well established that epigenetic marks play an important regulatory role in gene expression by modulating the interaction between transcriptional machinery and DNA regulatory elements commonly found in CpG islands and promoters [50–52]. While 5mC mapping to promoter regions is typically related to gene-expression repression [46], 5mC mapping to gene bodies is related to transcription activation [53]. 5hmC marks mapped to promoters and gene bodies are commonly found in actively transcribed genes [25]. 5hmC is involved in gene activation by increasing hydrophilicity, destabilizing the double helix and increasing interaction with transcription factors [54,55]. Notably, most of the identified GWS epigenetic marks in our study that map to genes enriched in mental disorders and PTSD showed predicted transcription-factor binding sites. This suggests that DNA 5mC/5hmC modifications in these disease-enriched genes may either hinder or enhance the binding of transcription factors to their sites, thereby influencing gene transcription [56]. Although our study did not allow for the direct examination of the transcriptional profile within the same OFC-neuronal nuclei, we complemented our neuro-specific findings by integrating them with bulk-transcriptomic results of the OFC reported in [9], where there is substantial sample overlap. We found convergence of five genes (*RTN4RL1, RASAL3, BIN2, FOXO3,* and *ITGAX*) across both-omics approaches. Future analysis employing single-cell epigenomic and transcriptomic approaches is warranted to gain a deeper understanding of the influence of differential 5mC/5hmC on gene expression patterns in the context of PTSD.

Our findings suggest that genes involved in cellular processes tend to be hyper-5mC in individuals with PTSD, while genes involved in embryonic morphogenesis and cell-fate commitment tend to be hyper-5hmC in individuals with PTSD. In the context of PTSD, it may result in a plausible downregulation of genes responsible for maintaining cell-cell connectivity, coupled with an upregulation of genes involved in cell-fate commitment. A recent study demonstrated that transcriptional dysregulation of genes involved in these two molecular mechanisms, particularly of 175 core genes, may disturb the homeostasis between neuronal survival and apoptosis throughout adulthood [57]. Of these 175 core genes, 12 were identified in our analyses: *RAB13, TWIST2, GABRA6* with differential 5mC and S*T14, PAQR5, MAF, TWIST2, PAQR5, PLEKHF1, TWIST2, OLIG2,* and *H1F0* with differential 5hmC. Considering our findings alongside the existing literature, we speculate that in the progression from trauma to PTSD, both 5mC and 5hmC may orchestrate a transcriptomic dysregulation in processes such as cell-cell connectivity and cell-fate commitment, disrupting the delicate equilibrium between neuronal survival and apoptosis.

A growing literature indicates that early life stress can significantly increase the risk of developing psychiatric disorders like PTSD by affecting brain development in key regions such as the prefrontal cortex, amygdala, and hippocampus, as well as by altering synaptic organization and stress response [58–62]. Animal models have shown alterations in spine density and dendritic length in the prefrontal cortex (including OFC) after stress exposure during different developmental stages [60,63–65]. Our study revealed a notable and consistent enrichment for developmental processes, as was evident in our GWS-differential 5mC/5hmC findings, co-5mC, co-5hmC network modules, and multi-omics convergence analysis of PTSD. Furthermore, we identified an enrichment of our GWS-differential 5hmC sites in early developmental stages including late fetal, early infancy, and early childhood. Notably, this enrichment, led by GWS- differential 5hmC sites, maps to genes such as *PAX, SOX, FOX, NOTCH,* and *HMX*, all key regulators of embryonic development of the central nervous system [66–68]. Overall, our findings suggest that the epigenetic dysregulation associated with PTSD, mainly 5hmC, might occur during early stages of development.

Studies examining genetic variants through GWAS, as well as epigenomic and transcriptomic profiles, have provided valuable insights into the genes and mechanisms associated with PTSD. However, most studies examine each-omic domain in isolation [7,69,70]. Cross-tissue multi-omics analysis may unveil converging molecular pathways involved in psychiatric disorders like PTSD while also shedding light on potential biomarkers and therapeutic targets [71,72]. In our integrative analysis, we delve into the intricate interplay of genetic, epigenetic, and transcriptomic factors across different tissues to reveal novel gene and gene targets for the treatment of PTSD. Among our most notable findings from the cross-tissue multi-omics analysis, we found genes participating in the hypothalamic-pituitary-adrenal axis (HPA) axis to be closely implicated in PTSD. Previous epigenetic studies in targeted genes have linked HPA-axis-related genes such as *NR3C1* and *FKBP5* with PTSD [73–77]. In the HPA axis, corticotropin-releasing hormone (CRH) orchestrates cortisol release upon exposure to stress. Cortisol then binds to the glucocorticoid receptor (*NR3C1*), along with other co-chaperones such as *FKBP5* and *HSP90*, to be translocated into the nucleus. This complex binds to glucocorticoid-response elements (GRE) in the promoter region of several genes to regulate their expression [78–80]. Corticotropin-releasing hormone receptor 1 (*CRHR1*), a well-known G-protein-coupled receptor that binds to CRH, has been identified in a recent large-scale GWAS of PTSD re-experiencing symptoms [81]. *CRHR1* was one of the identified genes with differential 5hmC at CpGs with a predicted TFBS. Further, our drug-repurposing analysis revealed the interaction of *CRHR1* with drugs such as verucerfont, ONO-2333MS, pexacerfont, SSR125543, ontamalimab, and antalarmin. Some of these drugs are currently being evaluated in clinical trials for the treatment of depression, including ONO-2333MS (NCT05923866) and SSR125543 (NCT01034995). Pexacerfont has been evaluated in humans as a potential treatment for drug dependence [82]. Antalarmin has not been evaluated in humans, but in animal models it has been shown to attenuate conditioned-fear response and reduce anxiety-associated behaviors [83,84]. In addition to the *CRHR1* gene, we found *HSP90AA1* and *BAIAP2* to be both differentially 5hmC and implicated in the HPA axis. *HSP90AA1* encodes a molecular chaperone that binds to *FKBP5* and mediates the nuclear translocation of glucocorticoid receptors [85,86]. *BAIAP2* (brain-specific angiogenesis inhibitor 1-associated protein 2) is a glucocorticoid-responsive protein involved in cytoskeletal reorganization related to membrane/actin dynamics at excitatory synapses and dendritic spine density in neurons. Future work is necessary to evaluate the viability of these genes as treatment targets for PTSD.

Other well-known biological pathways related to PTSD identified in our multi-omics convergent analysis include immune-system process, chemical-synaptic transmission, response to hormones, and the canonical Wnt/MAPK pathways. Glucocorticoids may affect both central and peripheral immune responses in PTSD, where increased production of pro-inflammatory cytokines may orchestrate neuroinflammation and neuron apoptosis [87–89]. The Wnt pathway, which regulates cell-fate specification and cell proliferation, has also been implicated in inflammation. For example, it interacts with NF-κB, which regulates the expression of immune-related genes such as cytokines, chemokines, and immune receptors [90]. Recently, upregulation of Wnt signaling was reported in peripheral blood mononuclear cells of individuals with PTSD, leading to an inflammatory phenotype [91].

Additional genes showing differential 5hmC at CpGs, with predicted TFBS and enrichment for mental disorders, as well as identified druggable targets in our drug repurposing analysis, include *DRD4* and *HDAC4.* Dopamine receptor 4 (*DRD4*), expressed in brain regions such as the hypothalamus, frontal cortex, and hippocampus, has been implicated in memory processes [92]. Polymorphisms identified in this gene have been associated with psychiatric disorders including PTSD [92–95]. The histone deacetylase 4 (*HDAC4*) gene is expressed in the amygdala and has been related to learning and memory [96]. An epigenome-wide association study found that methylation of *HDAC4* in blood is significantly associated with PTSD, and that differential expression of *HDAC4* in the amygdala of female rodents varies with estrogen levels [96]. An upregulation of *HDAC4* was also associated with PTSD symptoms in World Trade Center responders [97]. Additional convergent genes identified in our multi-omic analysis are *SOX6* and *NDRG1*, both reported in GWAS studies. A SNP mapping to an intron of N-myc downstream-regulated gene 1 (*NDRG1*), a stress-response gene that participates in processes such as hypoxia, inflammation, and cell growth, was reported to be significantly associated with PTSD [98]. The SNP rs12420134 mapping to *SOX6*, a key gene that participates in the development of the central nervous system, was significantly associated with hyper-arousal in PTSD individuals [8].

Functional implications of PTSD-associated GWS epigenetic marks at non-CpGs revealed genes involved in synaptic signaling such as *SHANK2*, *GABRA6* and *GRIN2C*, as well as those previously implicated in psychiatric disorders such as *HDAC4* and *GRIN1*. *SHANK2* encodes a scaffold protein in the postsynaptic density of excitatory synapses [99], *GRIN1* and *GRIN2C* encode critical subunits of N-methyl-D-aspartate (NMDA) receptors that regulate AMPAR-mediated excitatory and *GABRA6* encodes a subunits of gamma-aminobutyric acid type A (GABA) receptor, the major inhibitory neurotransmitter in the mammalian brain [100]. Exposure to chronic stress is linked to an excitatory-inhibitory (E/I) imbalance, including hypoactivity of the PFC due to an over-inhibition of GABAergic neurons [101,102]. Decreased levels of GABA in plasma is associated with development of PTSD [102]. Altogether, suggests that 5mC and 5hmC at non-CpGs may disrupt the excitatory-inhibitory (E/I) balance regulation in the OFC and play an important role in PTSD.

## Limitations

Considering the tissue and cell-type specificity of epigenetic patterns, future work should include a broader investigation across various neuronal types (glutamatergic and GABAergic) and extend to other brain cell types (oligodendrocytes, astrocytes, and microglia) as well as other brain regions (amygdala and hippocampus) implicated in the etiology of PTSD. Further, larger cohorts will allow for an evaluation of the effects of sex, age, and type of trauma on the epigenetic landscape in the human brain, which is essential to gaining a deeper understanding of the role of epigenetics in PTSD. By examining these diverse factors, we can achieve a more comprehensive and nuanced understanding of the complex interplay between epigenetics and the psychopathology of PTSD.

## Conclusion

In conclusion, our comprehensive multi-omic analysis assessing 5mC and 5hmC in both CpG and non-CpG sites represents a significant advance in our understanding of epigenetic contributions to PTSD. By integrating and evaluating the genomic distribution, target genes, and functional enrichment of 5mC and 5hmC, we provide insights into how each plays a distinct yet interconnected role in PTSD. In our multi-omics integrative analysis, we not only report molecular mechanisms and genes well known to be associated with PTSD, but also pinpoint potential therapeutic targets for effective treatment of PTSD. These approaches offer tangible avenues for future targeted intervention.

## Data Availability

All raw data produced in the present study are available upon reasonable request to the authors. Summary stats of the analysis will be publicly available in supplementary tables

## Acknowledgments

We would like to thank the brain donors and their families that made this research possible. We also thank the members of the Traumatic Stress Brain Research Group, as well as Sarah Beck for assisting with the English editing.

Members of the Traumatic Stress Brain Research Group include:

Victor E. Alvarez, MD^1^, David Benedek, MD^2^, Alicia Che, PhD^3^, Dianne A. Cruz, MS^4^, David A. Davis, PhD^5^, Matthew J. Girgenti, PhD^3,6^, Ellen Hoffman, MD, PhD^3^, Paul E. Holtzheimer, MD^6,7^, Bertrand R. Huber, MD, PhD^8^, Alfred Kaye, MD, PhD^3^, John H. Krystal, MD^3,6^, Adam T. Labadorf, PhD^8^, Terence M. Keane, PhD^6,8^, Mark W. Logue, PhD^6,8^, Ann McKee, MD^8^, Brian Marx, PhD^6,8^, Mark W. Miller, PhD^6,8^, Crystal Noller, PhD^6,7^, Janitza Montalvo-Ortiz, PhD^3^, William K. Scott, PhD^5^, Paula Schnurr, PhD^6,7^, Thor Stein, MD, PhD^8^, Robert Ursano, MD^7^, Douglas E. Williamson, PhD^4^, Erika J. Wolf, PhD^6,8^, Keith A. Young, PhD^9^.

Affiliations: 1. Boston University, Boston, MA. 2. Uniformed Services University of the Health Sciences, Bethesda, MD. 3. Yale University, New Haven, CT. 4. Duke University School of Medicine, Durham, NC. 5. University of Miami, Miami, FL. 6. National Center for PTSD. 7. Geisel School of Medicine at Dartmouth, Hanover, NH. 8. Boston University, Boston, MA. 9. Texas A&M University, College Station, TX.

## Funding Statement

This study was supported by the National Institute on Drug Abuse R21DA050160 and DP1DA058737 (J.L.M.O.); the U.S. Department of Veterans Affairs via the National Center for Posttraumatic Stress Disorder – Veterans Affairs Connecticut (J.L.M.O.), 1IK2CX002095-01A1 (J.L.M.O.), and the Kavli Institute for Neuroscience at Yale University Kavli Postdoctoral Award for Academic Diversity (J.J.M.M.). This publication was made possible in part by the Yale Center for Brain and Mind Health, which is sponsored by the Yale School of Medicine (D.L.N.R.).

## Author Contribution Statement

D.L.N.R. conducted the main analyses and drafted the manuscript; J.L.M.O. conceived, designed and coordinated the study, assisted in data interpretation, and revised the manuscript; Y.H. and G.R. were involved in data generation and interpretation and revised the manuscript; S.T.N. performed the co-methylation and co-hydroxymethylation analyses and assisted in manuscript writing; J.J.M.M. performed the drug-repurposing analysis and assisted in manuscript writing; and J.H.K. assisted in data interpretation, provided clinical feedback, and revised the manuscript. All authors contributed to and approved the final manuscript.

## Competing interests

John H. Krystal, MD served as a consultant for Aptinyx, Inc., Biogen, Idec, MA, Bionomics, Limited (Australia), Boehringer Ingelheim International, Epiodyne, Inc., EpiVario, Inc., Janssen Research & Development, Jazz Pharmaceuticals, Inc., Otsuka America Pharmaceutical, Inc., Spring Care, Inc., Sunovion Pharmaceuticals, Inc., Freedom Biosciences, Inc., Biohaven Pharmaceuticals, BioXcel Therapeutics, Inc. (Clinical Advisory Board), Cerevel Therapeutics, LLC, Delix Therapeutics, Inc., Eisai, Inc., EpiVario, Inc., Jazz Pharmaceuticals, Inc., Neumora Therapeutics, Inc., Neurocrine Biosciences, Inc., Novartis Pharmaceuticals Corporation, PsychoGenics, Inc., Takeda Pharmaceuticals, Tempero Bio, Inc., Terran Biosciences, Inc., Biohaven Pharmaceuticals, Freedom Biosciences, Spring Health, Inc., Biohaven Pharmaceuticals Medical Sciences, Cartego Therapeutics, Damona Pharmaceuticals, Delix Therapeutics, EpiVario, Inc., Neumora Therapeutics, Inc., Rest Therapeutics, Tempero Bio, Inc., Terran Biosciences, Inc., and Tetricus, Inc.

No additional authors report a conflict of interest.

## Data and materials availability

All data is available in supplementary files

## START Methods

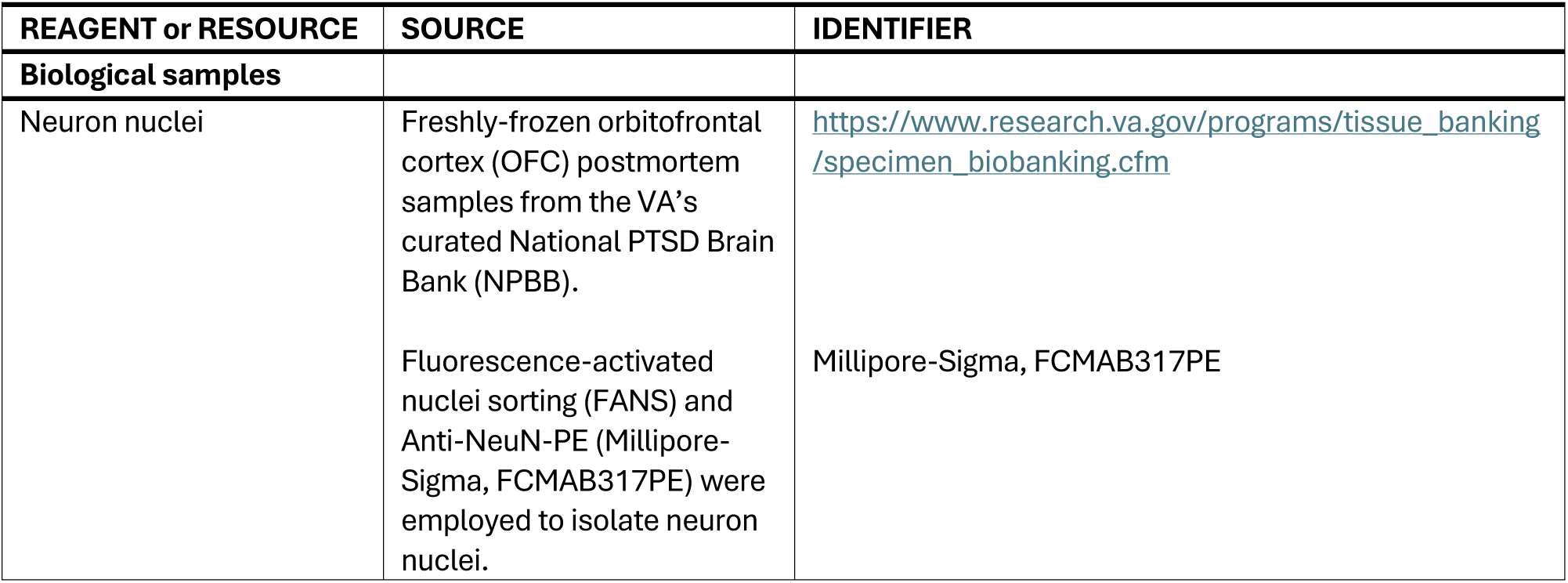
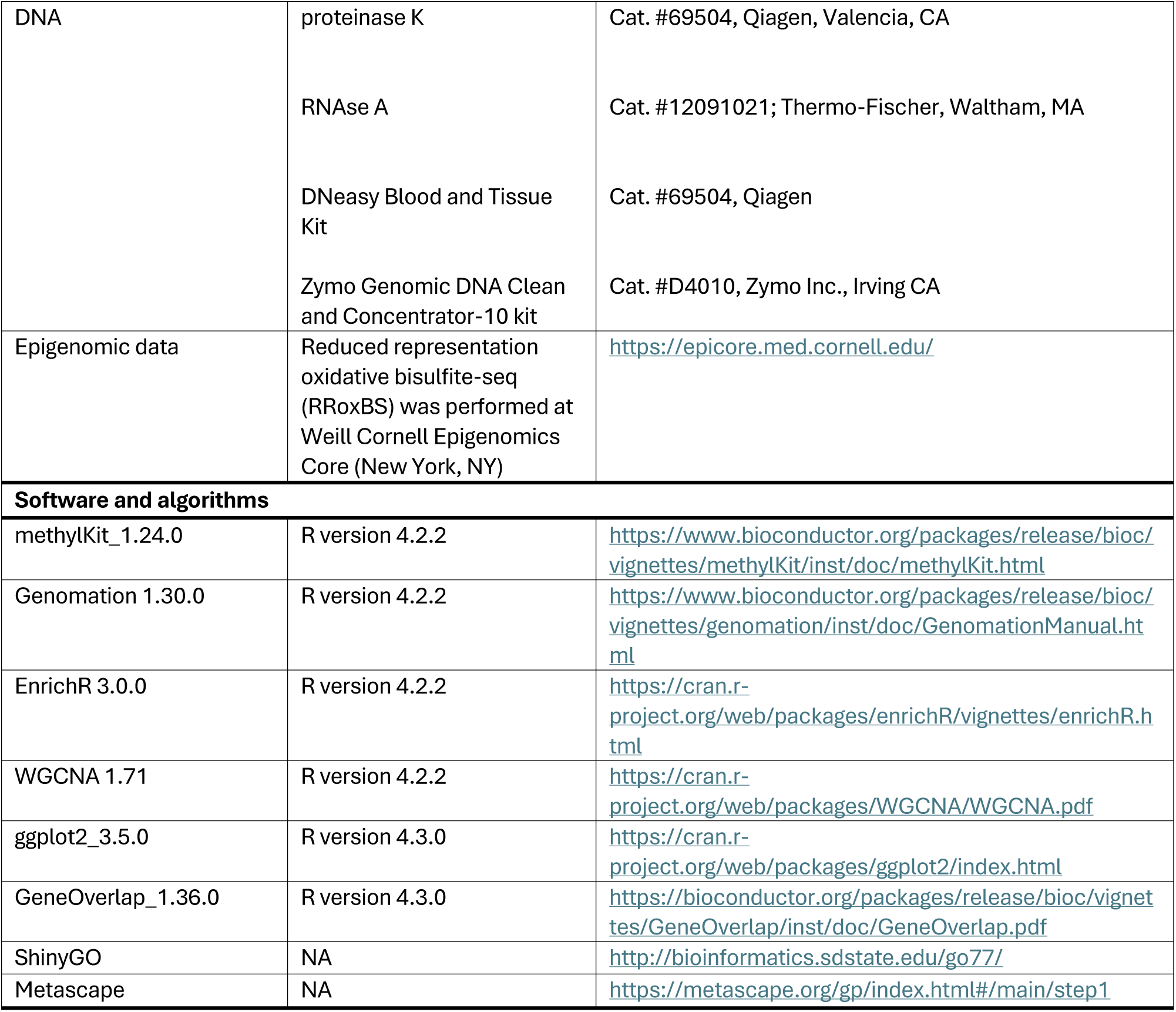

| REAGENT or RESOURCE | SOURCE | IDENTIFIER |
| --- | --- | --- |
| **Biological samples** |  |  |
| Neuron nuclei | Freshly-frozen orbitofrontal cortex (OFC) postmortem samples from the VA's curated National PTSD Brain Bank (NPBB).<br>Fluorescence-activated nuclei sorting (FANS) and Anti-NeuN-PE (Millipore-Sigma, FCMA317PE) were employed to isolate neuron nuclei. | https://www.research.va.gov/programs/tissue_banking/specimen_biobanking.cfm<br>Millipore-Sigma, FCMA317PE |
| DNA | proteinase K | Cat. #69504, Qiagen, Valencia, CA |
|  | RNAse A | Cat. #12091021; Thermo-Fischer, Waltham, MA |
|  | DNeasy Blood and Tissue Kit | Cat. #69504, Qiagen |
|  | Zymo Genomic DNA Clean and Concentrator-10 kit | Cat. #D4010, Zymo Inc., Irving CA |
| Epigenomic data | Reduced representation oxidative bisulfite-seq (RRoxBS) was performed at Weill Cornell Epigenomics Core (New York, NY) | https://epicore.med.cornell.edu/ |
| **Software and algorithms** |  |  |
| methyKit_1.24.0 | R version 4.2.2 | https://www.bioconductor.org/packages/release/bioc/vignettes/methyKit/inst/doc/methyKit.html |
| Genomation 1.30.0 | R version 4.2.2 | https://www.bioconductor.org/packages/release/bioc/vignettes/genomation/inst/doc/GenomationManual.html |
| EnrichR 3.0.0 | R version 4.2.2 | https://cran.r-project.org/web/packages/enrichR/vignettes/enrichR.html |
| WGCNA 1.71 | R version 4.2.2 | https://cran.r-project.org/web/packages/WGCNA/WGCNA.pdf |
| ggplot2_3.5.0 | R version 4.3.0 | https://cran.r-project.org/web/packages/ggplot2/index.html |
| GeneOverlap_1.36.0 | R version 4.3.0 | https://bioconductor.org/packages/release/bioc/vignettes/GeneOverlap/inst/doc/GeneOverlap.pdf |
| ShinyGO | NA | http://bioinformatics.sdstate.edu/go77/ |
| Metascape | NA | https://metascape.org/gp/index.html#/main/step1 |

## Materials and Methods

### Study cohort

Freshly-frozen orbitofrontal cortex (OFC) postmortem samples were obtained from the VA’s curated National PTSD Brain Bank (NPBB) [103], including 26 samples from individuals diagnosed with PTSD and 14 healthy controls. Detailed information on informed consent, diagnostic assessments, and brain tissue dissections can be found in [103]. Briefly, informed consent was obtained directly from the donors upon enrollment in NPBB or from their next of kin shortly after death. Clinical diagnosis under DSM-IV criteria was based on NPBB’s postmortem diagnostic assessment protocol, surveys, and medical record reviews. Each brain region was hemisected, cut into 1–1.5-cm slabs, and cooled on aluminum plates suspended in dry ice.

### Neuronal nuclei isolation and DNA extraction

For neuronal nuclei isolation, we conducted fluorescence-activated nuclei sorting (FANS) as previously described [33]. The OFC tissue was lysed, homogenized, filtered, and ultracentrifuged. The resulting neuronal nuclei were resuspended in 0.5% bovine serum albumin with Anti-NeuN-PE (Millipore-Sigma, FCMAB317PE). Next, we added 50 µL proteinase K (Cat. #69504, Qiagen, Valencia, CA) and 20 mg/mL RNAse A (Cat. #12091021; Thermo-Fischer, Waltham, MA) to the samples. Genomic DNA (gDNA) was isolated from the nuclei using the DNeasy Blood and Tissue Kit (Cat. #69504, Qiagen), following the manufacturer’s recommendations. The DNA samples were then concentrated using the Zymo Genomic DNA Clean and Concentrator-10 kit (Cat. #D4010, Zymo Inc., Irving CA) and stored at −80°C.

### High-throughput bisulfite sequencing and data processing

Reduced representation oxidative bisulfite-seq (RRoxBS) was carried out at the Weill Cornell Epigenomics Core (New York, NY). RRoxBS interrogates 5mC/5hmC in ∼85-90% of CpG islands [104]. Briefly, we used MspI digestion on 400 ng of gDNA to prepare the 5mC and 5hmC libraries with the TECAN Ovation RRoxBS Methyl-Seq library preparation kit. An oxidation step was performed only on the 5hmC library. Both libraries were then exposed to bisulfite conversion followed by a single-end 1 x 50 bp sequencing with the Illumina NovaSeq6000 system (mean depth of 42.7 +/- 1.5 (µ +/- SEM) million reads per library). The sequencing data underwent adapter trimming, alignment, and mapping-efficiency analysis using an in-house pipeline [105] at the Weill Cornell Epigenomics Core (New York, NY). The data was aligned and annotated to the human genome 38 (hg38).

### Differential 5mC/5hmC and enrichment analysis

Principal Component Analysis (PCA) identified two outliers that were removed from the analysis. The demographic and clinical information of the remaining 38 donors (25 PTSD cases and 13 healthy controls) is summarized in **Table S21**. The *MethylKit* R package [106] was utilized to identify differentially-methylated positions (DMP) or differentially-hydroxymethylated positions (DhMP) at CpGs and non-CpGs (CHG and CHH). For each analysis, sites were filtered based on coverage (>10x) and percentile of coverage in each sample (<99.9th). Normalization was carried out using the scaling factor derived from differences between mean or median of coverage distributions [106] to reduce bias in statistical tests. DMP and DhMP were identified at single-CpG/non-CpG resolution using the percentage of methylated and unmethylated cytosines. Logistic regression models were applied with overdispersion correction and *p*-values were adjusted using a sliding linear model (SLIM) to calculate *q*-values. Covariates such as ancestry, smoking, postmortem interval (PMI), and age of death (**Table S21**) were included in the model. GWS-differential 5mC or 5hmC cytosines were defined based on *q*-value < 0.05 and a differential 5mC/5hmC percentage greater than 2% between PTSD cases and controls (delta (Δ) value > 2%) [107–110]. We also assessed differentially-5mC/5hmC regions (DMR/DhMR) greater than 1000pb with at least 3 DMP/DhMP bases (cytosines) per region by using the tileMethylCounts function in the *methylkit* R package [106].

For genomic-feature annotation, we utilized the *Genomation* R package [111]. BED files of significant differentially 5mC / 5hmC sites were coerced into GRanges and then annotated with gene parts including promoters, introns, exons, and intergenic regions using the annotateWithGeneParts function. CpG-island annotation, including CpG islands, CpG island shores (∼2kb from islands), CpG shelves (∼4kb from islands), and open sea, was performed using the annotateWithFeatureFlank function. The distance from the nearest transcription start site (TSS) to DMP or DhMP was calculated using the getAssociationWithTSS function of genomation [111]. To refine DMP and DhMP mapping to gene-body regions, the nearest genes within 1.5Kb up- and downstream, and the nearest genes 10Kb up- and downstream, gene annotation was confirmed by using the UCSC Genome Browser annotation webtool [112]. CpGs farther than 10 kb from the nearest TSS were considered intergenic [113]. The JASPAR database [114,115], also integrated into the UCSC Genome Browser online resource [112], was used to identify predicted transcription-factor binding sites in the significant differential sites associated with PTSD.

Enrichment analysis was performed using the *enrichr* package in R [116,117], shinyGO [118], and the Metascape [119] online resources. This analysis incorporated gene names of the nearest transcription start site (TSS) located within a distance of < 10 kb from the genome-wide-significant (GWS) 5mC and 5hmC sites at CpGs and non-CpGs. Metascape integrates databases such as NCATS BioPlanet, Panther, Gene Ontology Consortium, and Kyoto Encyclopedia of Genes and Genomes (KEGG) [120]. Disease-enrichment analyses were conducted using DisGenet [121] integrated into Metascape [119]. Functional enrichments were considered significant with a corrected *p*-value (either FDR or *q*-value) < 0.05. Protein-protein interaction (PPI) analysis was carried out using the STRING online resource [122]. For the developmental-stages enrichment analysis, the Cell-type Specific Expression Analysis (CSEA) webtool (http://genetics.wustl.edu/jdlab/csea-tool-2/) was employed.

### Co-methylation analysis

Co-methylation analysis for 5mC and 5hmC at CpGs and non-CpGs was conducted using the *WGCNA* R package [123]. Significant modules (clusters of sites) were chosen as significant if they show correlations higher than 0.4 for CpGs and 0.2 for non-CpGs with PTSD, with a statistically significant p-value < 0.05. GO enrichment analysis was performed for each significant module using Metascape [119].

### Multi-omic integrative analysis

We assessed the convergence of our findings with those reported in the PTSD-omic literature examining human samples/tissue. The PTSD-omic studies evaluated included epigenomic studies in whole blood [13,124–129], transcriptomic studies from peripheral samples [57,130,139,131–138], transcriptomic studies of bulk OFC tissue [9], and GWAS studies of PTSD [7,8,81]. To test whether the convergence was statistically significant, we performed Fisher’s exact test employing the GeneOverlap R package [140]. To explore common molecular mechanisms underlying convergent PTSD-related findings, we used Metascape [119]. PTSD-associated differentially-expressed genes in the OFC were designated “DEG.OFC,” differentially-expressed genes in the peripheral tissue were called “Transcriptomic.Peripheral,” and differentially-methylated genes in the peripheral tissue were called “EWAS.” Genes identified in large-scale GWAS studies and those reported as associated with PTSD in the GWAS Catalog [141] were included in the “GWAS” category. Our PTSD-associated 5mC and 5hmC findings were categorized as “Neuro.5mC” and “Neuro.5hmC,” respectively. *P*-values of enriched terms were adjusted using Bonferroni correction in Metascape.

### Drug-repurposing analysis

For drug-repurposing analysis, we utilized the Drug Gene Interaction Database (DGIbd) [142], which analyzes drug-gene interactions and assigns an interaction score to prioritize gene-level data. We used a query score greater than or equal to 5 [142] to prioritize our findings.

## List of supplementary Materials

Table S1 Differentially methylated positions (DMP)

Table S2 Enrichment of differentially methylated positions (DMP)

Table S3 Enrichment of differentially methylated positions (DMP) by gene features

Table S4 Top significant enriched pathways by co-methylated modules

Table S5 Differentially hydroxymethylated positions (DhMP)

Table S6 Enrichment of differentially hydroxymethylated positions (DhMP)

Table S7 Enrichment of differentially hydroxymethylated positions (DhMP) by gene features

Table S8 Top significant enriched pathways by co-hydroxymethylated modules

Table S9 Convergent pathways of DMP and DhMP

Table S10 Convergent pathways of DMP and DhMP per cytosines distribution

Table S11 Brain region and developmental stage enrichment DMP

Table S12 Brain region and developmental stage enrichment DhMP

Table S13 Disease enrichment analysis for DMP

Table S14 Disease enrichment analysis for DhMP

Table S15 Convergent disease enrichment analysis for DMP and DhMP

Table S16 Convergent disease enrichment analysis for DMP and DhMP by epigenetic mark

Table S17 Muilt-omic convergence

Table S18 Convergent enrichment analysis of OFC findings

Table S19 Convergent enrichment analysis of multi-omics findings

Table S20 Drug discovery analysis

Table S21 Demographic and clinical information of the study cohort

Fig S1. Functional enrichment analysis for both GWS 5mC CpGs and non-CpGs. Fig S2. Co-methylation analysis

Fig S3. Functional enrichment analysis for both GWS 5hmC CpGs and non-CpGs Fig S4. Co-hydroxymethylation analysis

Fig S5. Convergent functional enrichment analysis of GWS-differential 5mC and 5hmC CpG and non-CpG sites.

Fig S6. Convergent functional enrichment analysis of GWS-differential 5mC and 5hmC CpG and non-CpG sites along with GWS-differentially-expressed genes in the bulk-OFC

Fig S7. Convergent functional enrichment analysis of GWS-differential 5mC and 5hmC CpG and non-CpG sites along with other omics findings.

## Notes

### Author Declarations

Samples used here were collected from the National Posttraumatic Stress Disorder Brain Bank, which is part of the US Department of Veterans Affairs (VA). As part of the VA, any local VA facility to be a participating site in a research project that is reviewed by the VA Central IRB, the local facility must also enter into a Memorandum of Understanding (MOU) with the VHA Central Office Human Research Protections Program (HRPP), and rewrite its SOPs to incorporate the processes for using the VA CIRB. Both ORO and ORD must approve changes in the facilities IRB relationships, so the MOU will be reviewed by each. I confirm that all necessary patient/participant consent has been obtained and the appropriate institutional forms have been archived, and that any patient/participant/sample identifiers included were not known to anyone (e.g., hospital staff, patients or participants themselves) outside the research group so cannot be used to identify individuals.

## References

[1] APA. American Psychiatric Association, 2013. Diagnostic and statistical manual of mental disorders (5th ed.). 2013.

[2] Brady KT, Back SE. Childhood Trauma, Posttraumatic Stress Disorder, and Alcohol Dependence. Alcohol Res 2012;34:408.

[3] AS Z, N P, EB B. Epigenetics of Posttraumatic Stress Disorder: Current Evidence, Challenges, and Future Directions. Biol Psychiatry 2015;78:327–35. 10.1016/J.BIOPSYCH.2015.04.003.

[4] GS T, BJ D, KR D, Y Z, M R-F, R H, et al. Systems biology approach to understanding post-traumatic stress disorder. Mol Biosyst 2015;11:980–93. 10.1039/C4MB00404C.

[5] Lee J-Y, Kim S-W, Kim J-M. The Impact of Community Disaster Trauma: A Focus on Emerging Research of PTSD and Other Mental Health Outcomes. Chonnam Med J 2020;56. 10.4068/cmj.2020.56.2.99.

[6] Kilpatrick DG, Resnick HS, Milanak ME, Miller MW, Keyes KM, Friedman MJ. National Estimates of Exposure to Traumatic Events and PTSD Prevalence Using DSM-IV and DSM-5 Criteria. J Trauma Stress 2013;26. 10.1002/jts.21848.

[7] Nievergelt CM, Maihofer AX, Klengel T, Atkinson EG, Chen C-Y, Choi KW, et al. International meta-analysis of PTSD genome-wide association studies identifies sex- and ancestry-specific genetic risk loci. Nat Commun 2019 101 2019;10:1–16. 10.1038/s41467-019-12576-w.

[8] Stein MB, Levey DF, Cheng Z, Wendt FR, Harrington K, Pathak GA, et al. Genome-wide association analyses of post-traumatic stress disorder and its symptom subdomains in the Million Veteran Program. Nat Genet 2021 532 2021;53:174–84. 10.1038/s41588-020-00767-x.

[9] Girgenti MJ, Wang J, Ji D, Cruz DA, Stein MB, Gelernter J, et al. Transcriptomic organization of the human brain in post-traumatic stress disorder. Nat Neurosci 2020 241 2020;24:24–33. 10.1038/s41593-020-00748-7.

[10] Smith ZD, Meissner A. DNA methylation: Roles in mammalian development. Nat Rev Genet 2013;14. 10.1038/nrg3354.

[11] Hammamieh R, Chakraborty N, Gautam A, Muhie S, Yang R, Donohue D, et al. Whole-genome DNA methylation status associated with clinical PTSD measures of OIF/OEF veterans. Transl Psychiatry 2017;7. 10.1038/tp.2017.129.

[12] Mehta D, Klengel T, Conneely KN, Smith AK, Altmann A, Pace TW, et al. Childhood maltreatment is associated with distinct genomic and epigenetic profiles in posttraumatic stress disorder. Proc Natl Acad Sci 2013;110:8302–7. 10.1073/PNAS.1217750110.

[13] Montalvo-Ortiz JL, Gelernter J, Cheng Z, Girgenti MJ, Xu K, Zhang X, et al. Epigenome-wide association study of posttraumatic stress disorder identifies novel loci in U.S. military veterans. Transl Psychiatry 2022;12:65. 10.1038/S41398-022-01822-3.

[14] Morrison FG, Miller MW, Logue MW, Assef M, Wolf EJ. DNA Methylation Correlates of PTSD: Recent Findings and Technical Challenges. Prog Neuropsychopharmacol Biol Psychiatry 2019;90:223. 10.1016/J.PNPBP.2018.11.011.

[15] Rutten BPF, Vermetten E, Vinkers CH, Ursini G, Daskalakis NP, Pishva E, et al. Longitudinal analyses of the DNA methylome in deployed military servicemen identify susceptibility loci for post-traumatic stress disorder. Mol Psychiatry 2018;23. 10.1038/mp.2017.120.

[16] Smith AK, Ratanatharathorn A, Maihofer AX, Naviaux RK, Aiello AE, Amstadter AB, et al. Epigenome-wide meta-analysis of PTSD across 10 military and civilian cohorts identifies novel methylation loci. BioRxiv 2019. 10.1101/585109.

[17] Muhie S, Gautam A, Meyerhoff J, Chakraborty N, Hammamieh R, Jett M. Brain transcriptome profiles in mouse model simulating features of post-traumatic stress disorder. Mol Brain 2015;8. 10.1186/S13041-015-0104-3.

[18] Muhie S, Gautam A, Chakraborty N, Hoke A, Meyerhoff J, Hammamieh R, et al. Molecular indicators of stress-induced neuroinflammation in a mouse model simulating features of post-traumatic stress disorder. Transl Psychiatry 2017;7:e1135. 10.1038/TP.2017.91.

[19] Núñez-Rios DL, Martínez-Magaña JJ, Nagamatsu ST, Krystal JH, Martínez-González KG, Giusti-Rodríguez P, et al. Cross-Species Convergence of Brain Transcriptomic and Epigenomic Findings in Posttraumatic Stress Disorder: A Systematic Review. Complex Psychiatry 2023;9. 10.1159/000529536.

[20] Li D, Yang Y, Li Y, Zhu X, Li Z. Epigenetic regulation of gene expression in response to environmental exposures: From bench to model. Sci Total Environ 2021;776. 10.1016/j.scitotenv.2021.145998.

[21] Hahn MA, Qiu R, Wu X, Li AX, Zhang H, Wang J, et al. Dynamics of 5-Hydroxymethylcytosine and Chromatin Marks in Mammalian Neurogenesis. Cell Rep 2013;3. 10.1016/j.celrep.2013.01.011.

[22] Irwin RE, Thakur A, O’ Neill KM, Walsh CP. 5-Hydroxymethylation marks a class of neuronal gene regulated by intragenic methylcytosine levels. Genomics 2014;104. 10.1016/j.ygeno.2014.08.013.

[23] Khare T, Pai S, Koncevicius K, Pal M, Kriukiene E, Liutkeviciute Z, et al. 5-hmC in the brain is abundant in synaptic genes and shows differences at the exon-intron boundary. Nat Struct Mol Biol 2012;19. 10.1038/nsmb.2372.

[24] Szulwach KE, Li X, Li Y, Song CX, Han JW, Kim SS, et al. Integrating 5-hydroxymethylcytosine into the epigenomic landscape of human embryonic stem cells. PLoS Genet 2011;7. 10.1371/journal.pgen.1002154.

[25] Bachman M, Uribe-Lewis S, Yang X, Williams M, Murrell A, Balasubramanian S. 5- Hydroxymethylcytosine is a predominantly stable DNA modification. Nat Chem 2014;6. 10.1038/nchem.2064.

[26] Johnson KC, Houseman EA, King JE, Von Herrmann KM, Fadul CE, Christensen BC. 5- Hydroxymethylcytosine localizes to enhancer elements and is associated with survival in glioblastoma patients. Nat Commun 2016;7. 10.1038/ncomms13177.

[27] Stroud H, Feng S, Morey Kinney S, Pradhan S, Jacobsen SE. 5-Hydroxymethylcytosine is associated with enhancers and gene bodies in human embryonic stem cells. Genome Biol 2011;12. 10.1186/gb-2011-12-6-r54.

[28] Marion-Poll L, Roussarie JP, Taing L, Dard-Dascot C, Servant N, Jaszczyszyn Y, et al. DNA methylation and hydroxymethylation characterize the identity of D1 and D2 striatal projection neurons. Commun Biol 2022;5. 10.1038/s42003-022-04269-w.

[29] Spruijt CG, Gnerlich F, Smits AH, Pfaffeneder T, Jansen PWTC, Bauer C, et al. Dynamic readers for 5-(Hydroxy)methylcytosine and its oxidized derivatives. Cell 2013;152. 10.1016/j.cell.2013.02.004.

[30] Gross JA, Pacis A, Chen GG, Drupals M, Lutz PE, Barreiro LB, et al. Gene-body 5- hydroxymethylation is associated with gene expression changes in the prefrontal cortex of depressed individuals. Transl Psychiatry 2017;7. 10.1038/tp.2017.93.

[31] Clark SL, Chan RF, Zhao M, Xie LY, Copeland WE, Penninx BWJH, et al. Dual methylation and hydroxymethylation study of alcohol use disorder. Addict Biol 2021. 10.1111/adb.13114.

[32] Andrade-Brito DE, Núñez-Ríos DL, Martínez-Magaña JJ, Nagamatsu ST, Rompala G, Zillich L, et al. Neuronal-specific methylome and hydroxymethylome analysis reveal significant loci associated with alcohol use disorder. Front Genet 2024;15. 10.3389/fgene.2024.1345410.

[33] Rompala G, Nagamatsu ST, Martínez-Magaña JJ, Nuñez-Ríos DL, Wang J, Girgenti MJ, et al. Profiling neuronal methylome and hydroxymethylome of opioid use disorder in the human orbitofrontal cortex. Nat Commun 2023;14:4544. 10.1038/s41467-023-40285-y.

[34] de Mendoza A, Poppe D, Buckberry S, Pflueger J, Albertin CB, Daish T, et al. The emergence of the brain non-CpG methylation system in vertebrates. Nat Ecol Evol 2021;5. 10.1038/s41559-020-01371-2.

[35] Jang HS, Shin WJ, Lee JE, Do JT. CpG and non-CpG methylation in epigenetic gene regulation and brain function. Genes (Basel) 2017;8. 10.3390/genes8060148.

[36] Kozlenkov A, Roussos P, Timashpolsky A, Barbu M, Rudchenko S, Bibikova M, et al. Differences in DNA methylation between human neuronal and glial cells are concentrated in enhancers and non-CpG sites. Nucleic Acids Res 2014;42. 10.1093/nar/gkt838.

[37] Lister R, Pelizzola M, Dowen RH, Hawkins RD, Hon G, Tonti-Filippini J, et al. Human DNA methylomes at base resolution show widespread epigenomic differences. Nature 2009;462. 10.1038/nature08514.

[38] Schultz MD, He Y, Whitaker JW, Hariharan M, Mukamel EA, Leung D, et al. Human body epigenome maps reveal noncanonical DNA methylation variation. Nature 2015;523. 10.1038/nature14465.

[39] Nagamatsu ST, Rompala G, Hurd YL, Núñez-Rios DL, Montalvo-Ortiz JL, Alvarez VE, et al. CpH methylome analysis in human cortical neurons identifies novel gene pathways and drug targets for opioid use disorder. Front Psychiatry 2023;13. 10.3389/fpsyt.2022.1078894.

[40] Bremner JD, Vythilingam M, Vermetten E, Southwick SM, McGlashan T, Staib LH, et al. Neural correlates of declarative memory for emotionally valenced words in women with posttraumatic stress disorder related to early childhood sexual abuse. Biol Psychiatry 2003;53. 10.1016/S0006-3223(02)01891-7.

[41] Jackowski AP, de Araújo Filho GM, de Almeida AG, de Araújo CM, Reis M, Nery F, et al. The involvement of the orbitofrontal cortex in psychiatric disorders: an update of neuroimaging findings. Rev Bras Psiquiatr 2012;34. 10.1590/s1516-44462012000200014.

[42] Shin LM, McNally RJ, Kosslyn SM, Thompson WL, Rauch SL, Alpert NM, et al. Regional cerebral blood flow during script-driven imagery in childhood sexual abuse-related PTSD: A PET investigation. Am J Psychiatry 1999;156. 10.1176/ajp.156.4.575.

[43] Shin LM, Orr SP, Carson MA, Rauch SL, Macklin ML, Lasko NB, et al. Regional Cerebral Blood Flow in the Amygdala and Medial Prefrontal Cortex during Traumatic Imagery in Male and Female Vietnam Veterans with PTSD. Arch Gen Psychiatry 2004;61. 10.1001/archpsyc.61.2.168.

[44] Long HK, King HW, Patient RK, Odom DT, Klose RJ. Protection of CpG islands from DNA methylation is DNA-encoded and evolutionarily conserved. Nucleic Acids Res 2016;44. 10.1093/nar/gkw258.

[45] Sharma RP, Gavin DP, Grayson DR. CpG methylation in neurons: Message, memory, or mask. Neuropsychopharmacology 2010;35. 10.1038/npp.2010.85.

[46] Jin SG, Wu X, Li AX, Pfeifer GP. Genomic mapping of 5-hydroxymethylcytosine in the human brain. Nucleic Acids Res 2011;39. 10.1093/nar/gkr120.

[47] Schlosberg CE, Wu DY, Gabel HW, Edwards JR. ME-Class2 reveals context dependent regulatory roles for 5-hydroxymethylcytosine. Nucleic Acids Res 2019;47. 10.1093/nar/gkz001.

[48] He Y, Ecker JR. Non-CG Methylation in the Human Genome. Annu Rev Genomics Hum Genet 2015;16. 10.1146/annurev-genom-090413-025437.

[49] Lee JH, Saito Y, Park SJ, Nakai K. Existence and possible roles of independent non-CpG methylation in the mammalian brain. DNA Res 2021;27. 10.1093/DNARES/DSAA020.

[50] Bird AP. CpG islands as gene markers in the vertebrate nucleus. Trends Genet 1987;3. 10.1016/0168-9525(87)90294-0.

[51] Hughes AL, Kelley JR, Klose RJ. Understanding the interplay between CpG island-associated gene promoters and H3K4 methylation. Biochim Biophys Acta - Gene Regul Mech 2020;1863. 10.1016/j.bbagrm.2020.194567.

[52] Le NQK, Yapp EKY, Nagasundaram N, Yeh HY. Classifying Promoters by Interpreting the Hidden Information of DNA Sequences via Deep Learning and Combination of Continuous FastText N-Grams. Front Bioeng Biotechnol 2019;7. 10.3389/fbioe.2019.00305.

[53] Breiling A, Lyko F. Epigenetic regulatory functions of DNA modifications: 5-methylcytosine and beyond. Epigenetics and Chromatin 2015;8. 10.1186/s13072-015-0016-6.

[54] Ehrlich M, Ehrlich KC. DNA cytosine methylation and hydroxymethylation at the borders. Epigenomics 2014;6. 10.2217/epi.14.48.

[55] Yu M, Hon GC, Szulwach KE, Song CX, Zhang L, Kim A, et al. Base-resolution analysis of 5- hydroxymethylcytosine in the mammalian genome. Cell 2012;149. 10.1016/j.cell.2012.04.027.

[56] Brochet P, Ianni BM, Laugier L, Frade AF, Silva Nunes JP, Teixeira PC, et al. Epigenetic regulation of transcription factor binding motifs promotes Th1 response in Chagas disease cardiomyopathy. Front Immunol 2022;13. 10.3389/fimmu.2022.958200.

[57] Morello G, Villari A, Spampinato AG, La Cognata V, Guarnaccia M, Gentile G, et al. Transcriptional profiles of cell fate transitions reveal early drivers of neuronal apoptosis and survival. Cells 2021;10. 10.3390/cells10113238.

[58] Nie Y, Wen L, Song J, Wang N, Huang L, Gao L, et al. Emerging trends in epigenetic and childhood trauma: Bibliometrics and visual analysis. Front Psychiatry 2022;13. 10.3389/fpsyt.2022.925273.

[59] Chen Y, Baram TZ. Toward understanding how early-life stress reprograms cognitive and emotional brain networks. Neuropsychopharmacology 2016;41. 10.1038/npp.2015.181.

[60] Kolb B, Gibb R. Brain Plasticity and Behaviour in the Developing Brain. J Can Acad Child Adolesc Psychiatry 2011;20.

[61] Pervanidou P, Makris G, Chrousos G, Agorastos A. Early life stress and pediatric posttraumatic stress disorder. Brain Sci 2020;10. 10.3390/brainsci10030169.

[62] Smith KE, Pollak SD. Early life stress and development: potential mechanisms for adverse outcomes. J Neurodev Disord 2020;12. 10.1186/s11689-020-09337-y.

[63] Muhammad A, Hossain S, Pellis SM, Kolb B. Tactile Stimulation During Development Attenuates Amphetamine Sensitization and Structurally Reorganizes Prefrontal Cortex and Striatum in a Sex-Dependent Manner. Behav Neurosci 2011;125. 10.1037/a0022628.

[64] Murmu MS, Salomon S, Biala Y, Weinstock M, Braun K, Bock J. Changes of spine density and dendritic complexity in the prefrontal cortex in offspring of mothers exposed to stress during pregnancy. Eur J Neurosci 2006;24. 10.1111/j.1460-9568.2006.05024.x.

[65] Mychasiuk R, Gibb R, Kolb B. Prenatal bystander stress induces neuroanatomical changes in the prefrontal cortex and hippocampus of developing rat offspring. Brain Res 2011;1412. 10.1016/j.brainres.2011.07.023.

[66] Angelozzi M, Lefebvre V. SOXopathies: Growing Family of Developmental Disorders Due to SOX Mutations. Trends Genet 2019;35. 10.1016/j.tig.2019.06.003.

[67] Hannenhalli S, Kaestner KH. The evolution of Fox genes and their role in development and disease. Nat Rev Genet 2009;10. 10.1038/nrg2523.

[68] Prior HM, Walter MA. Sox genes: Architects of development. Mol Med 1996;2. 10.1007/bf03401900.

[69] Uddin M, Ratanatharathorn A, Armstrong D, Kuan PF, Aiello AE, Bromet EJ, et al. Epigenetic meta-analysis across three civilian cohorts identifies NRG1 and HGS as blood-based biomarkers for post-traumatic stress disorder. Epigenomics 2018;10. 10.2217/epi-2018-0049.

[70] Kuan PF, Waszczuk MA, Kotov R, Clouston S, Yang X, Singh PK, et al. Gene expression associated with PTSD in World Trade Center responders: An RNA sequencing study. Transl Psychiatry 2017;7. 10.1038/s41398-017-0050-1.

[71] Guan F, Ni T, Zhu W, Williams LK, Cui LB, Li M, et al. Integrative omics of schizophrenia: from genetic determinants to clinical classification and risk prediction. Mol Psychiatry 2021. 10.1038/s41380-021-01201-2.

[72] Cong S, Sang Z, Cao L, Yuan J, Li Y, Liang H, et al. Integration of tissue-specific multi-omics data implicates brain targets for complex neuropsychiatric traits. MedRxiv 2023:2023.06.14.23291366. 10.1101/2023.06.14.23291366.

[73] Alexander N, Kirschbaum C, Wankerl M, Stauch BJ, Stalder T, Steudte-Schmiedgen S, et al. Glucocorticoid receptor gene methylation moderates the association of childhood trauma and cortisol stress reactivity. Psychoneuroendocrinology 2018;90. 10.1016/j.psyneuen.2018.01.020.

[74] Bustamante AC, Aiello AE, Galea S, Ratanatharathorn A, Noronha C, Wildman DE, et al. Glucocorticoid receptor DNA methylation, childhood maltreatment and major depression. J Affect Disord 2016;206. 10.1016/j.jad.2016.07.038.

[75] Klengel T, Mehta D, Anacker C, Rex-Haffner M, Pruessner JC, Pariante CM, et al. Allele-specific FKBP5 DNA demethylation mediates gene-childhood trauma interactions. Nat Neurosci 2013;16:33–41. 10.1038/NN.3275.

[76] Palma-Gudiel H, Córdova-Palomera A, Leza JC, Fañanás L. Glucocorticoid receptor gene (NR3C1) methylation processes as mediators of early adversity in stress-related disorders causality: A critical review. Neurosci Biobehav Rev 2015;55:520–35. 10.1016/J.NEUBIOREV.2015.05.016.

[77] Wilker S, Pfeiffer A, Kolassa S, Elbert T, Lingenfelder B, Ovuga E, et al. The role of FKBP5 genotype in moderating long-term effectiveness of exposure-based psychotherapy for posttraumatic stress disorder. Transl Psychiatry 2014;4. 10.1038/TP.2014.49.

[78] Dunlop BW, Wong A. The hypothalamic-pituitary-adrenal axis in PTSD: Pathophysiology and treatment interventions. Prog Neuro-Psychopharmacology Biol Psychiatry 2019;89:361–79. 10.1016/J.PNPBP.2018.10.010.

[79] Girgenti MJ, Duman RS. Transcriptome Alterations in Posttraumatic Stress Disorder. Biol Psychiatry 2018;83:840–8. 10.1016/J.BIOPSYCH.2017.09.023.

[80] Nishi M. Effects of Early-Life Stress on the Brain and Behaviors: Implications of Early Maternal Separation in Rodents. Int J Mol Sci 2020;21. 10.3390/IJMS21197212.

[81] Gelernter J, Sun N, Polimanti R, Pietrzak R, Levey DF, Bryois J, et al. Genome-wide association study of post-traumatic stress disorder reexperiencing symptoms in >165,000 US veterans. Nat Neurosci 2019;22:1394–401. 10.1038/s41593-019-0447-7.

[82] Morabbi MJ, Razaghi E, Moazen-Zadeh E, Safi-Aghdam H, Zarrindast MR, Vousoghi N, et al. Pexacerfont as a CRF1 antagonist for the treatment of withdrawal symptoms in men with heroin/methamphetamine dependence: A randomized, double-blind, placebo-controlled clinical trial. Int Clin Psychopharmacol 2018;33. 10.1097/YIC.0000000000000200.

[83] Deak T. The Impact of the Nonpeptide Corticotropin-Releasing Hormone Antagonist Antalarmin on Behavioral and Endocrine Responses to Stress. Endocrinology 1999;140. 10.1210/en.140.1.79.

[84] Cursano S, Battaglia CR, Urrutia-Ruiz C, Grabrucker S, Schön M, Bockmann J, et al. A CRHR1 antagonist prevents synaptic loss and memory deficits in a trauma-induced delirium-like syndrome. Mol Psychiatry 2021;26. 10.1038/s41380-020-0659-y.

[85] Flasbeck V, Brüne M. Association between childhood maltreatment, psychopathology and DNA methylation of genes involved in stress regulation: Evidence from a study in Borderline Personality Disorder. PLoS One 2021;16. 10.1371/JOURNAL.PONE.0248514.

[86] Kirschke E, Goswami D, Southworth D, Griffin PR, Agard DA. Glucocorticoid Receptor Function Regulated by Coordinated Action of the Hsp90 and Hsp70 Chaperone Cycles. Cell 2014;157:1685–97. 10.1016/J.CELL.2014.04.038.

[87] Silverman MN, Sternberg EM. Glucocorticoid regulation of inflammation and its behavioural and metabolic correlates: from HPA axis to glucocorticoid receptor dysfunction. Ann N Y Acad Sci 2012;1261.

[88] Kertser A, Baruch K, Deczkowska A, Weiner A, Croese T, Kenigsbuch M, et al. Corticosteroid signaling at the brain-immune interface impedes coping with severe psychological stress. Sci Adv 2019;5. 10.1126/sciadv.aav4111.

[89] Merlo D, Cuchillo-Ibañez I, Parlato R, Rammes G. DNA Damage, Neurodegeneration, and Synaptic Plasticity. Neural Plast 2016;2016. 10.1155/2016/1206840.

[90] Jridi I, Canté-Barrett K, Pike-Overzet K, Staal FJT. Inflammation and Wnt Signaling: Target for Immunomodulatory Therapy? Front Cell Dev Biol 2021;8. 10.3389/fcell.2020.615131.

[91] Bam M, Yang X, Busbee BP, Aiello AE, Uddin M, Ginsberg JP, et al. Increased H3K4me3 methylation and decreased miR-7113-5p expression lead to enhanced Wnt/β-catenin signaling in immune cells from PTSD patients leading to inflammatory phenotype. Mol Med 2020;26:1–15. 10.1186/S10020-020-00238-3/FIGURES/6.

[92] Magwai T, Xulu KR. Physiological Genomics Plays a Crucial Role in Response to Stressful Life Events, the Development of Aggressive Behaviours, and Post-Traumatic Stress Disorder (PTSD). Genes (Basel) 2022;13. 10.3390/genes13020300.

[93] Blum K, Gondré-Lewis MC, Modestino EJ, Lott L, Baron D, Siwicki D, et al. Understanding the Scientific Basis of Post-traumatic Stress Disorder (PTSD): Precision Behavioral Management Overrides Stigmatization. Mol Neurobiol 2019;56. 10.1007/s12035-019-1600-8.

[94] Hoxha B, Uka AG, Agani F, Haxhibeqiri S, Haxhibeqiri V, Dzananovic ES, et al. The role of TAQI DRD2 (rs1800497) and DRD4 VNTR polymorphisms in posttraumatic stress disorder (PTSD). Psychiatr Danub 2019;31. 10.24869/psyd.2019.263.

[95] Sánchez-Mora C, Richarte V, Garcia-Martínez I, Pagerols M, Corrales M, Bosch R, et al. Dopamine receptor DRD4 gene and stressful life events in persistent attention deficit hyperactivity disorder. Am J Med Genet Part B Neuropsychiatr Genet 2015;168. 10.1002/ajmg.b.32340.

[96] Maddox SA, Kilaru V, Shin J, Jovanovic T, Almli LM, Dias BG, et al. Estrogen-dependent association of HDAC4 with fear in female mice and women with PTSD. Mol Psychiatry 2018 233 2017;23:658–65. 10.1038/mp.2016.250.

[97] Marchese S, Cancelmo L, Diab O, Cahn L, Aaronson C, Daskalakis NP, et al. Altered gene expression and PTSD symptom dimensions in World Trade Center responders. Mol Psychiatry 2022;27. 10.1038/s41380-022-01457-2.

[98] Xie P, Kranzler HR, Yang C, Zhao H, Farrer LA, Gelernter J. Genome-wide association study identifies new susceptibility loci for posttraumatic stress disorder. Biol Psychiatry 2013;74. 10.1016/j.biopsych.2013.04.013.

[99] Yoo YE, Yoo T, Kang H, Kim E. Brain region and gene dosage-differential transcriptomic changes in Shank2-mutant mice. Front Mol Neurosci 2022;15. 10.3389/fnmol.2022.977305.

[100] Zhou L, Sun X, Duan J. NMDARs regulate the excitatory-inhibitory balance within neural circuits. Brain Sci Adv 2023;9. 10.26599/bsa.2022.9050020.

[101] Page CE, Coutellier L. Prefrontal excitatory/inhibitory balance in stress and emotional disorders: Evidence for over-inhibition. Neurosci Biobehav Rev 2019;105. 10.1016/j.neubiorev.2019.07.024.

[102] Huang J, Xu F, Yang L, Tuolihong L, Wang X, Du Z, et al. Involvement of the GABAergic system in PTSD and its therapeutic significance. Front Mol Neurosci 2023;16. 10.3389/fnmol.2023.1052288.

[103] Friedman MJ, Huber BR, Brady CB, Ursano RJ, Benedek DM, Kowall NW, et al. VA’s National PTSD Brain Bank: a National Resource for Research. Curr Psychiatry Rep 2017;19. 10.1007/s11920-017-0822-6.

[104] Seiler Vellame D, Castanho I, Dahir A, Mill J, Hannon E. Characterizing the properties of bisulfite sequencing data: maximizing power and sensitivity to identify between-group differences in DNA methylation. BMC Genomics 2021;22. 10.1186/s12864-021-07721-z.

[105] Garrett-Bakelman FE, Sheridan CK, Kacmarczyk TJ, Ishii J, Betel D, Alonso A, et al. Enhanced reduced representation bisulfite sequencing for assessment of DNA methylation at base pair resolution. J Vis Exp 2015. 10.3791/52246.

[106] Akalin A, Kormaksson M, Li S, Garrett-Bakelman FE, Figueroa ME, Melnick A, et al. MethylKit: a comprehensive R package for the analysis of genome-wide DNA methylation profiles. Genome Biol 2012;13. 10.1186/gb-2012-13-10-R87.

[107] Ebbert MTW, Ross CA, Pregent LJ, Lank RJ, Zhang C, Katzman RB, et al. Conserved DNA methylation combined with differential frontal cortex and cerebellar expression distinguishes C9orf72-associated and sporadic ALS, and implicates SERPINA1 in disease. Acta Neuropathol 2017;134. 10.1007/s00401-017-1760-4.

[108] Gunawardhana LP, Baines KJ, Mattes J, Murphy VE, Simpson JL, Gibson PG. Differential DNA methylation profiles of infants exposed to maternal asthma during pregnancy. Pediatr Pulmonol 2014;49. 10.1002/ppul.22930.

[109] Legendre C, Gooden GC, Johnson K, Martinez RA, Liang WS, Salhia B. Whole-genome bisulfite sequencing of cell-free DNA identifies signature associated with metastatic breast cancer. Clin Epigenetics 2015;7. 10.1186/s13148-015-0135-8.

[110] Yue W, Cheng W, Liu Z, Tang Y, Lu T, Zhang D, et al. Genome-wide DNA methylation analysis in obsessive-compulsive disorder patients. Sci Rep 2016;6. 10.1038/srep31333.

[111] Akalin A, Franke V, Vlahoviček K, Mason CE, Schübeler D. Genomation: A toolkit to summarize, annotate and visualize genomic intervals. Bioinformatics 2015;31. 10.1093/bioinformatics/btu775.

[112] Kuhn RM, Haussler D, James Kent W. The UCSC genome browser and associated tools. Brief Bioinform 2013;14. 10.1093/bib/bbs038.

[113] Slieker RC, Bos SD, Goeman JJ, Bovée JV, Talens RP, Van Der Breggen R, et al. Identification and systematic annotation of tissue-specific differentially methylated regions using the Illumina 450k array. Epigenetics and Chromatin 2013;6. 10.1186/1756-8935-6-26.

[114] Bryne JC, Valen E, Tang MHE, Marstrand T, Winther O, Da piedade I, et al. JASPAR, the open access database of transcription factor-binding profiles: New content and tools in the 2008 update. Nucleic Acids Res 2008;36. 10.1093/nar/gkm955.

[115] Fornes O, Castro-Mondragon JA, Khan A, Van Der Lee R, Zhang X, Richmond PA, et al. JASPAR 2020: Update of the open-Access database of transcription factor binding profiles. Nucleic Acids Res 2020;48. 10.1093/nar/gkz1001.

[116] Jawaid W. enrichR: Provides an R Interface to “Enrichr.” R Packag Version 21 2019.

[117] Kuleshov M V., Jones MR, Rouillard AD, Fernandez NF, Duan Q, Wang Z, et al. Enrichr: a comprehensive gene set enrichment analysis web server 2016 update. Nucleic Acids Res 2016;44. 10.1093/nar/gkw377.

[118] Ge SX, Jung D, Jung D, Yao R. ShinyGO: A graphical gene-set enrichment tool for animals and plants. Bioinformatics 2020;36. 10.1093/bioinformatics/btz931.

[119] Zhou Y, Zhou B, Pache L, Chang M, Khodabakhshi AH, Tanaseichuk O, et al. Metascape provides a biologist-oriented resource for the analysis of systems-level datasets. Nat Commun 2019 101 2019;10:1–10. 10.1038/s41467-019-09234-6.

[120] Kanehisa M, Goto S. KEGG: Kyoto Encyclopedia of Genes and Genomes. Nucleic Acids Res 2000;28. 10.1093/nar/28.1.27.

[121] Piñero J, Ramírez-Anguita JM, Saüch-Pitarch J, Ronzano F, Centeno E, Sanz F, et al. The DisGeNET knowledge platform for disease genomics: 2019 update. Nucleic Acids Res 2020;48. 10.1093/nar/gkz1021.

[122] Szklarczyk D, Gable AL, Lyon D, Junge A, Wyder S, Huerta-Cepas J, et al. STRING v11: protein–protein association networks with increased coverage, supporting functional discovery in genome-wide experimental datasets. Nucleic Acids Res 2019;47:D607–13. 10.1093/NAR/GKY1131.

[123] Langfelder P, Horvath S. WGCNA: An R package for weighted correlation network analysis. BMC Bioinformatics 2008;9. 10.1186/1471-2105-9-559.

[124] Katrinli S, Maihofer AX, Wani AH, Pfeiffer JR, Ketema E, Ratanatharathorn A, et al. Epigenome-wide meta-analysis of PTSD symptom severity in three military cohorts implicates DNA methylation changes in genes involved in immune system and oxidative stress n.d.;14:17. 10.1038/s41380-021-01398-2.

[125] Kuan PF, Waszczuk MA, Kotov R, Marsit CJ, Guffanti G, Gonzalez A, et al. An epigenome-wide DNA methylation study of PTSD and depression in World Trade Center responders. Transl Psychiatry 2017;7. 10.1038/tp.2017.130.

[126] Logue MW, Miller MW, Wolf EJ, Huber BR, Morrison FG, Zhou Z, et al. An epigenome-wide association study of posttraumatic stress disorder in US veterans implicates several new DNA methylation loci. Clin Epigenetics 2020;12. 10.1186/s13148-020-0820-0.

[127] Ratanatharathorn A, Boks MP, Maihofer AX, Aiello AE, Amstadter AB, Ashley-Koch AE, et al. Epigenome-wide association of PTSD from heterogeneous cohorts with a common multi-site analysis pipeline. Am J Med Genet Part B Neuropsychiatr Genet 2017;174. 10.1002/ajmg.b.32568.

[128] Smith AK, Ratanatharathorn A, Maihofer AX, Naviaux RK, Aiello AE, Amstadter AB, et al. Epigenome-wide meta-analysis of PTSD across 10 military and civilian cohorts identifies methylation changes in AHRR. Nat Commun 2020;11. 10.1038/s41467-020-19615-x.

[129] Snijders C, Maihofer AX, Ratanatharathorn A, Baker DG, Boks MP, Geuze E, et al. Longitudinal epigenome-wide association studies of three male military cohorts reveal multiple CpG sites associated with post-traumatic stress disorder. Clin Epigenetics 2020;12. 10.1186/s13148-019-0798-7.

[130] Bhatt S, Hillmer AT, Girgenti MJ, Rusowicz A, Kapinos M, Nabulsi N, et al. PTSD is associated with neuroimmune suppression: evidence from PET imaging and postmortem transcriptomic studies. Nat Commun 2020;11. 10.1038/S41467-020-15930-5.

[131] Breen MS, Tylee DS, Maihofer AX, Neylan TC, Mehta D, Binder EB, et al. PTSD Blood Transcriptome Mega-Analysis: Shared Inflammatory Pathways across Biological Sex and Modes of Trauma. Neuropsychopharmacology 2018;43. 10.1038/npp.2017.220.

[132] Gautam A, D’Arpa P, Donohue DE, Muhie S, Chakraborty N, Luke BT, et al. Acute and chronic plasma metabolomic and liver transcriptomic stress effects in a mouse model with features of post-traumatic stress disorder. PLoS One 2015;10. 10.1371/journal.pone.0117092.

[133] Hori H, Yoshida F, Itoh M, Lin M, Niwa M, Ino K, et al. Proinflammatory status-stratified blood transcriptome profiling of civilian women with PTSD. Psychoneuroendocrinology 2020;111. 10.1016/j.psyneuen.2019.104491.

[134] Jeanneteau F, Barrè C, Vos M, De Vries CJM, Rouillard X, Levesque D, et al. The Stress-Induced Transcription Factor NR4A1 Adjusts Mitochondrial Function and Synapse Number in Prefrontal Cortex. J Neurosci 2018;38:1335–50. 10.1523/JNEUROSCI.2793-17.2017.

[135] Kuan PF, Ren X, Clouston S, Yang X, Jonas K, Kotov R, et al. PTSD is associated with accelerated transcriptional aging in World Trade Center responders. Transl Psychiatry 2021;11. 10.1038/s41398-021-01437-0.

[136] Lori A, Maddox SA, Sharma S, Andero R, Ressler KJ, Smith AK. Dynamic patterns of threat-associated gene expression in the amygdala and blood. Front Psychiatry 2019;10:778.

[137] Mehta D, Voisey J, Bruenig D, Harvey W, Morris CP, Lawford B, et al. Transcriptome analysis reveals novel genes and immune networks dysregulated in veterans with PTSD. Brain Behav Immun 2018;74. 10.1016/j.bbi.2018.08.014.

[138] Rusch HL, Robinson J, Yun S, Osier ND, Martin C, Brewin CR, et al. Gene expression differences in PTSD are uniquely related to the intrusion symptom cluster: A transcriptome-wide analysis in military service members. Brain Behav Immun 2019;80. 10.1016/j.bbi.2019.04.039.

[139] Torshizi AD, Wang K. Deconvolution of Transcriptional Networks in Post-Traumatic Stress Disorder Uncovers Master Regulators Driving Innate Immune System Function. Sci Rep 2017;7. 10.1038/s41598-017-15221-y.

[140] Shen L. GeneOverlap : An R package to test and visualize gene overlaps. Http://Shenlab-SinaiGithubIo/Shenlab-Sinai/2014.

[141] Sollis E, Mosaku A, Abid A, Buniello A, Cerezo M, Gil L, et al. The NHGRI-EBI GWAS Catalog: knowledgebase and deposition resource. Nucleic Acids Res 2023;51. 10.1093/nar/gkac1010.

[142] Freshour SL, Kiwala S, Cotto KC, Coffman AC, McMichael JF, Song JJ, et al. Integration of the Drug-Gene Interaction Database (DGIdb 4.0) with open crowdsource efforts. Nucleic Acids Res 2021;49:D1144–51. 10.1093/NAR/GKAA1084.

